# STRchive: a dynamic resource detailing population-level and locus-specific insights at tandem repeat disease loci

**DOI:** 10.1101/2024.05.21.24307682

**Authors:** Laurel Hiatt, Ben Weisburd, Egor Dolzhenko, Grace E. VanNoy, Edibe Nehir Kurtas, Heidi L. Rehm, Aaron Quinlan, Harriet Dashnow

## Abstract

Approximately 3% of the human genome consists of repetitive elements called tandem repeats (TRs), which include short tandem repeats (STRs) of 1–6bp motifs and variable number tandem repeats (VNTRs) of 7+bp motifs. TR variants contribute to several dozen mono- and polygenic diseases but remain understudied and “enigmatic,” particularly relative to single nucleotide variants. It remains comparatively challenging to interpret the clinical significance of TR variants. Although existing resources provide portions of necessary data for interpretation at disease-associated loci, it is currently difficult or impossible to efficiently invoke the additional details critical to proper interpretation, such as motif pathogenicity, disease penetrance, and age of onset distributions. It is also often unclear how to apply population information to analyses.

We present STRchive (S-T-archive, http://strchive.org/), a dynamic resource consolidating information on TR disease loci in humans from research literature, up-to-date clinical resources, and large-scale genomic databases, with the goal of streamlining TR variant interpretation at disease-associated loci. We apply STRchive —including pathogenic thresholds, motif classification, and clinical phenotypes—to a gnomAD cohort of ∼18.5k individuals genotyped at 60 disease-associated loci.

Through detailed literature curation, we demonstrate that the majority of TR diseases affect children despite being thought of as adult diseases. Additionally, we show that pathogenic genotypes can be found within gnomAD which do not necessarily overlap with known disease prevalence, and leverage STRchive to interpret locus-specific findings therein. We apply a diagnostic blueprint empowered by STRchive to relevant clinical vignettes, highlighting possible pitfalls in TR variant interpretation. As a living resource, STRchive is maintained by experts, takes community contributions, and will evolve as understanding of TR diseases progresses.

## Introduction

Tandem repeats (TRs) include short tandem repeats (STRs, 1–6 base pair motifs) and variable number tandem repeats (VNTRs, motifs of 7+ base pairs). These two highly mutable classes combined comprise approximately 3% of the human genome and cause numerous human diseases.^1–5^ STRs alone contribute to dozens of polygenic and monogenic diseases, with more than 60 Mendelian diseases caused by STR expansions.^6,7^ These STR conditions are estimated to collectively affect 1 in 3,000 people, with most disease burden presumed to be in undiagnosed individuals.^8^

The propensity of TR diseases to remain undiagnosed reflects the unique challenges of TR variant detection and interpretation. TRs remain understudied and “enigmatic,”^1^ particularly when compared to single nucleotide variants (SNVs). Long-standing difficulties analyzing repetitive sequences stem from mappability issues inherent to these low-complexity genomic regions.^9^ TRs thus have been historically overlooked due to technical challenges in genotyping, even after the advent of next-generation sequencing.^10,11^ Short-read sequencing remains problematic because TRs often approach or exceed the length of the read.^12,13^ While long-read sequencing offers technical improvements through expanded read length, obstacles to genotyping include stutter “noise” from polymerase during sequencing, or a distribution of allele sizes around the original allele, and low coverage leading to limited read support.^14^ Consequently, TRs are often excluded from routine genetic studies, or only well-established loci are considered.^14,15^ As TRs are thought to address some of the “missing heritability” in genetic disease, their continued absence in research and clinical efforts is a major shortcoming.^16,17^ As stated by Treangen and Salzberg, “simply ignoring repeats is not an option.”^18^

However, even when TRs are included in genetic assays, interpretation of variants remains difficult. Established filtering strategies—such as leveraging inheritance patterns, sequencing depth, and presumed functional impact^12,19^—can empower some interpretation, but the added complexity of TRs challenges many filtering norms. While these variants exist within the coding space of the genome, filtering to these regions risks missing TRs with potential functional impact in non-coding regions. Population frequency metrics based on hundreds of thousands of individuals in resources such as gnomAD and TOPMed enable the identification of rare SNV variants, which are more likely to be associated with disease.^19^ However, normal repeat ranges for TRs are often inferred by family studies or control cohorts that are several times smaller than the cohorts used in SNV analyses.^20^ Additionally, TRs are exceptionally polymorphic, with 10–10,000-fold higher mutation rates than non-repetitive loci.^5^ This extensive mutability can further exacerbate ancestry-specific allelic distributions,^15,21,22^ and large-scale allele frequency distributions are typically unavailable outside well-studied disease loci.^20^ Furthermore, most loci are described in European cohorts or small families during disease discovery without capturing the full extent of allellic diversity.^23^ Intermediate alleles, or premutations, may correspond to mild, preclinical, or variable phenotypes, such with Fragile X syndrome (FXS) versus late-onset Fragile X-associated tremor/ataxia syndrome (FXTAS).^9,24^ but many loci have intermediate allele size ranges for which pathogenicity is ambiguous or unknown due to a paucity of observations. Consequently, the threshold at which TR pathogenicity occurs is frequently unclear and subject to ongoing investigation.^9^

These genetic, phenotypic, and diagnostic complexities necessitate the cataloging of TR locus features for diagnostic and research purposes, and efforts have been made as the field develops.^12^ A subset of TR diseases are documented in the Clinical Genome Resource^25^ and associated variant database ClinVar^26^, particularly diseases localized to coding regions. However, the extent of TR-specific documentation is inconsistent and report-dependent, with diagnostic criteria generally unavailable in these resources.

GeneReviews offers clinically relevant peer-reviewed information on thousands of genetic conditions—including many TR diseases—but there is a delay from discovery to database inclusion that can last years, and reports differ substantially in detail by disease.^27^ Online Mendelian Inheritance in Man (OMIM) has a broadly consistent level of detail for each phenotype-gene relationship; however, its records encompass all variant types rather than providing TR-specific information, and its comprehensive reports can be difficult to parse into discrete, actionable details.^28^ None of these tools centralize TR disease loci into a single navigable repository, which is a major strength of the STRipy STRs database and the Genome Aggregation Database (gnomAD) table of TR disease loci.^29,30^ These resources currently include 65 and 60 loci, respectively, with documentation for reference region, canonical repeat motif, and—for most loci—normal versus pathogenic allele ranges.

Additionally, both databases have population-level allele distributions stratified by ancestry (2.5k individuals and five ancestry groups in STRipy; 18.5k and ten groups in gnomAD). gnomAD also provides the additional granularity of sex, genotyped motif, and, in some cases, sample age. Still, neither STRipy nor gnomAD capture the full information necessary for TR variant interpretation, such as age of symptom onset, estimated disease prevalence, and theorized pathogenicmechanisms.

We present STRchive (S-T-archive, http://strchive.org/), a dynamic resource that consolidates information on TR disease loci in humans from current literature, up-to-date research findings, and large-scale genomic databases. We combine automated pipelines for literature management with expert curation to ensure currency and accuracy within STRchive. As a comprehensive and version-controlled database, STRchive can empower diagnostic efforts and TR research initiatives.^15,16^ Crucially, we interpret the allelic distributions and genotype frequencies in ∼18.5k TR disease-unaffected individuals from gnomAD v3.1.3 in the wider context of disease prevalence, clinical phenotype, and diagnostic factors, as distilled within STRchive.

## Results

STRchive 1.0.0 contains aggregate information on 68 disease-associated loci, including 64 STR and four VNTR disease loci, drawn from the literature—including primary reports, case studies, and reviews—and major genomic resources such as OMIM and GeneReviews. Key citations are included within the database, and comprehensive locus-specific literature is cataloged and available to STRchive users. Disease loci were selected based on multiple instances of evidence across the literature and clinical genetics databases, with the first iteration of loci selection conducted on TR review papers^1,7^ and GeneReviews. These loci were then cross-referenced with the Tandem Repeats Finder track in the UCSC Genome Browser to establish a reference region and locus details were then augmented by relevant literature.^31,32^ STRchive is available as a user-friendly website and in machine-readable CSV and JSON formats for integration into variant calling and analysis pipelines. GRCh37, GRCh38, and CHM13-T2T coordinates are reported for each locus.

Within these 68 loci, preliminary loci with less published evidence are annotated with qualifiers, as are conflicting and refuted loci. Links to locus-specific pages in resources such as OMIM, GeneReviews, gnomAD, and STRipy are provided where available.

### STRchive combines automatic and supervised curation for comprehensive cataloging

We devised an automated PubMed search query (**Supplementary Methods**) to systematically and routinely update our database with locus-specific literature. This automated pipeline is run routinely and additions to locus literature are evaluated by our team of contributors for integration into STRchive. This approach is complemented by ongoing manual literature curation and community contributions of current and novel loci evaluated by the STRchive team. Resultantly, we catalog extensive information for each disease-associated locus, from genomic location and motif length to allele size ranges as relevant to pathogenicity (**Figure 1**).

**Figure 1:**
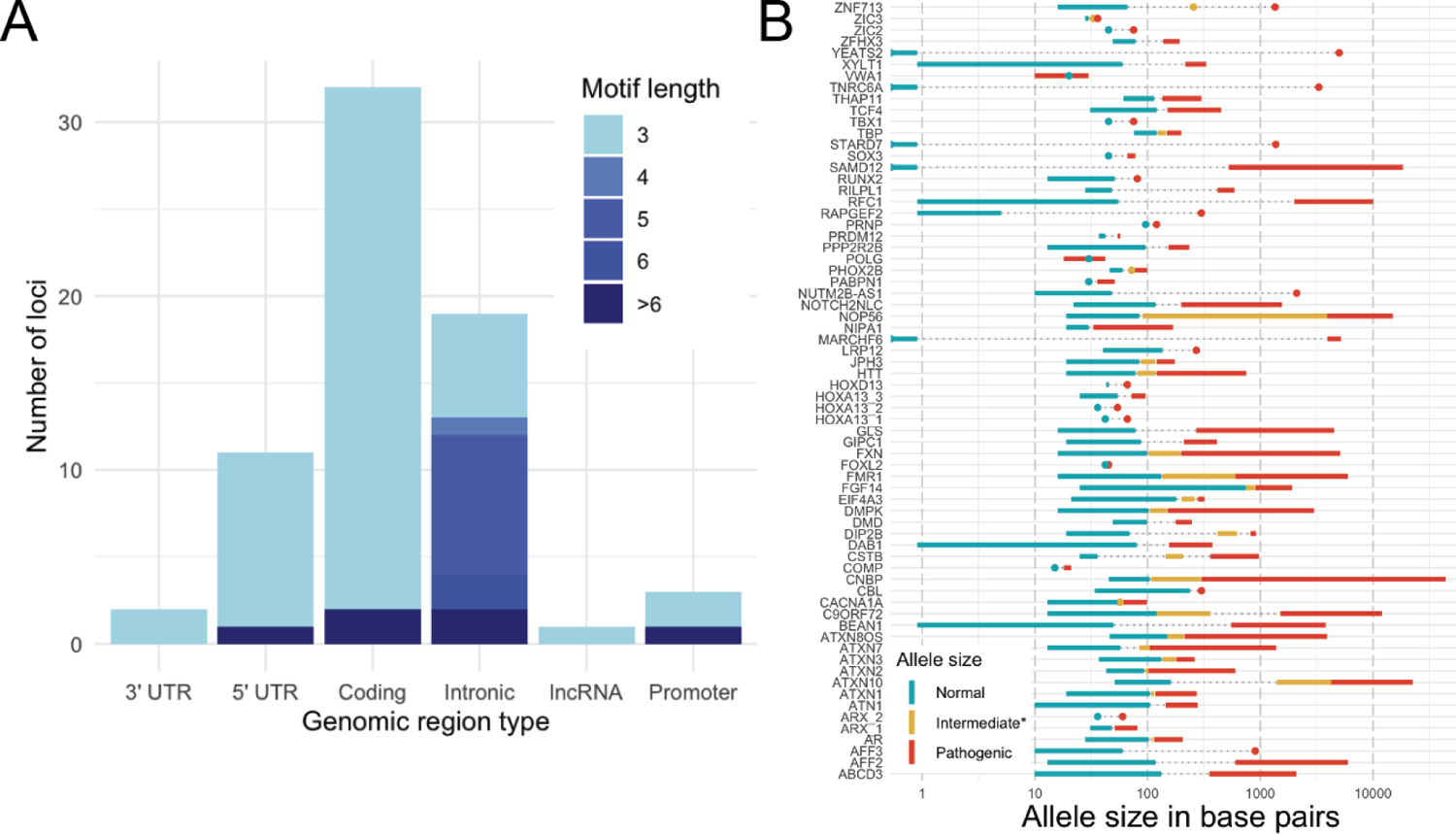
STRchive documents essential information across TR disease loci, from sequence context to locus-specific data. **A.** TR locus counts by motif size and genomic context. **B.** Ranges of literature-established allele sizes in bp. The intermediate size range indicates either a premutation, incomplete penetrance or an uncertain threshold of pathogenicity; circles indicate a value rather than an interval. Where there are no intermediate values but pathogenic thresholds are greater than the upper limit of the normal thresholds, dashed gray lines have been added. Independent observations, defined as unrelated cases/pedigrees as documented in OMIM, GeneReviews, and research literature; loci with less than two independent observations, or unrelated clinical cases, were removed, as was *POLG* (see **Methods**).

From our automated literature retrieval, we identify the earliest PubMed-indexed publication indicating the discovery of an associated monogenic disease at a TR locus. We contrast the number of unique PubMed IDs (PMIDs) including and after the earliest publication with the number of independent observations (or non-related clinical cases) supporting the disease association, manually curated from the literature (Figure 2).^1^ These publications were identified by explicit PubMed queries mentioning tandem repeats, human disease, and the locus gene (**Methods**). We capture the trend of increased discovery of TR loci as in the past decade as parallels advances in molecular and bioinformatic methods.^33^

**Figure 2:**
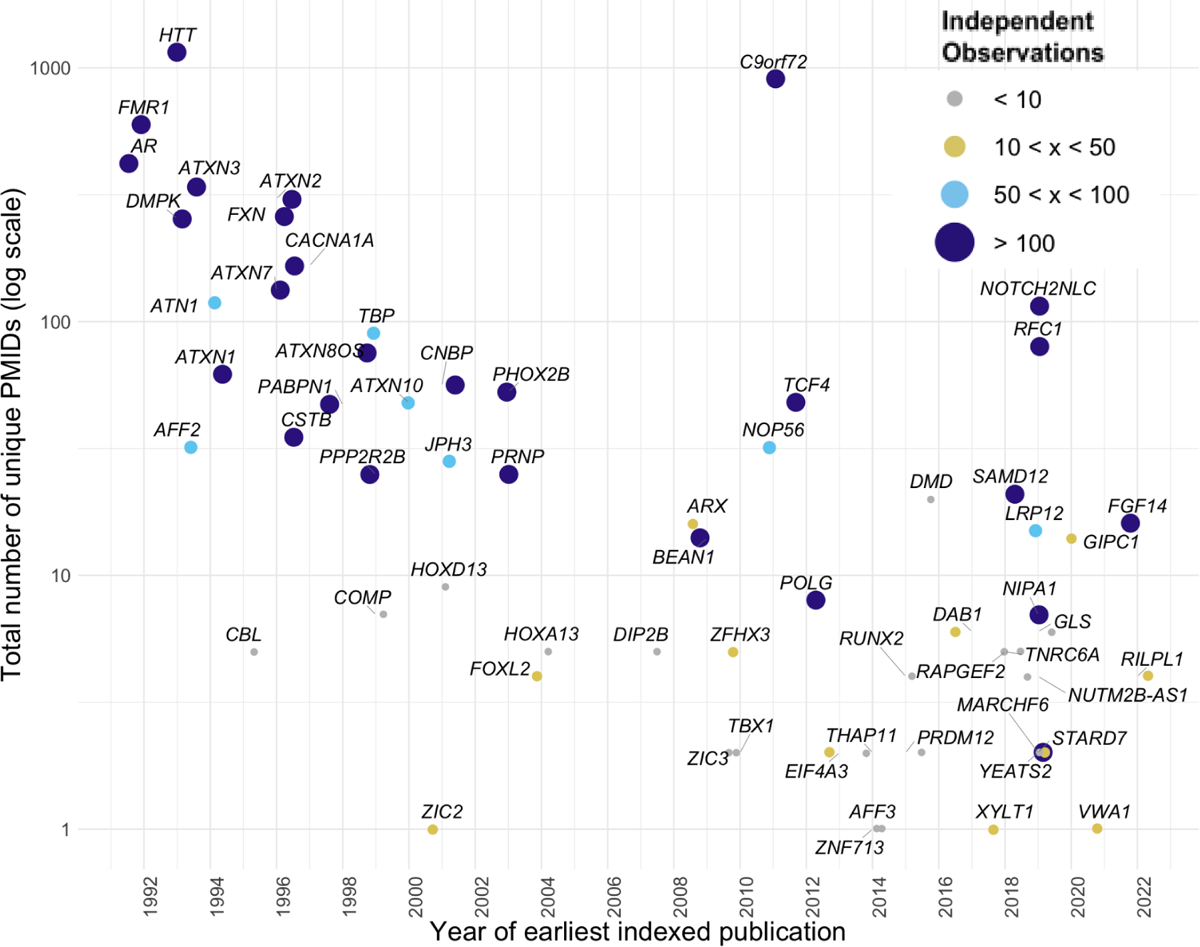
Locus-specific data, from literature catalog to clinical evidence, are captured by automated and manual curation. Total number of PMIDs with available PubMed year of discovery or earliest mention in indexed literature (as of April 30, 2024). Loci are colored and sized by the number of independent observations, defined as unrelated cases/pedigrees as documented in OMIM, GeneReviews, and research literature. Jitter is used to separate data points; years are considered as whole integers.

### STRchive reveals potential for childhood onset for a majority of TR diseases

While TR diseases are often thought to primarily affect adults due to allele instability over the lifetime,^34^ 82% (56/68) of documented TR conditions can affect children, with a documented case under the age of 18. Over a third (24/68) can present in the first year of life. To our knowledge, this is the first instance in which sufficient data have been aggregated to challenge the dogma of TR diseases as specific to adults. To determine whether pediatric cases fall within the expected range of disease onset or exist as outliers, we annotate the evidence supporting each loci and assign literature-based typical onset ranges where there are ten or more independent observations (Figure 3). We observe wide ranges of disease onset for well-documented diseases: the higher the prevalence and penetrance of a disease, the more likely we are to observe age variation due to a greater extent of case documentation.

**Figure 3:**
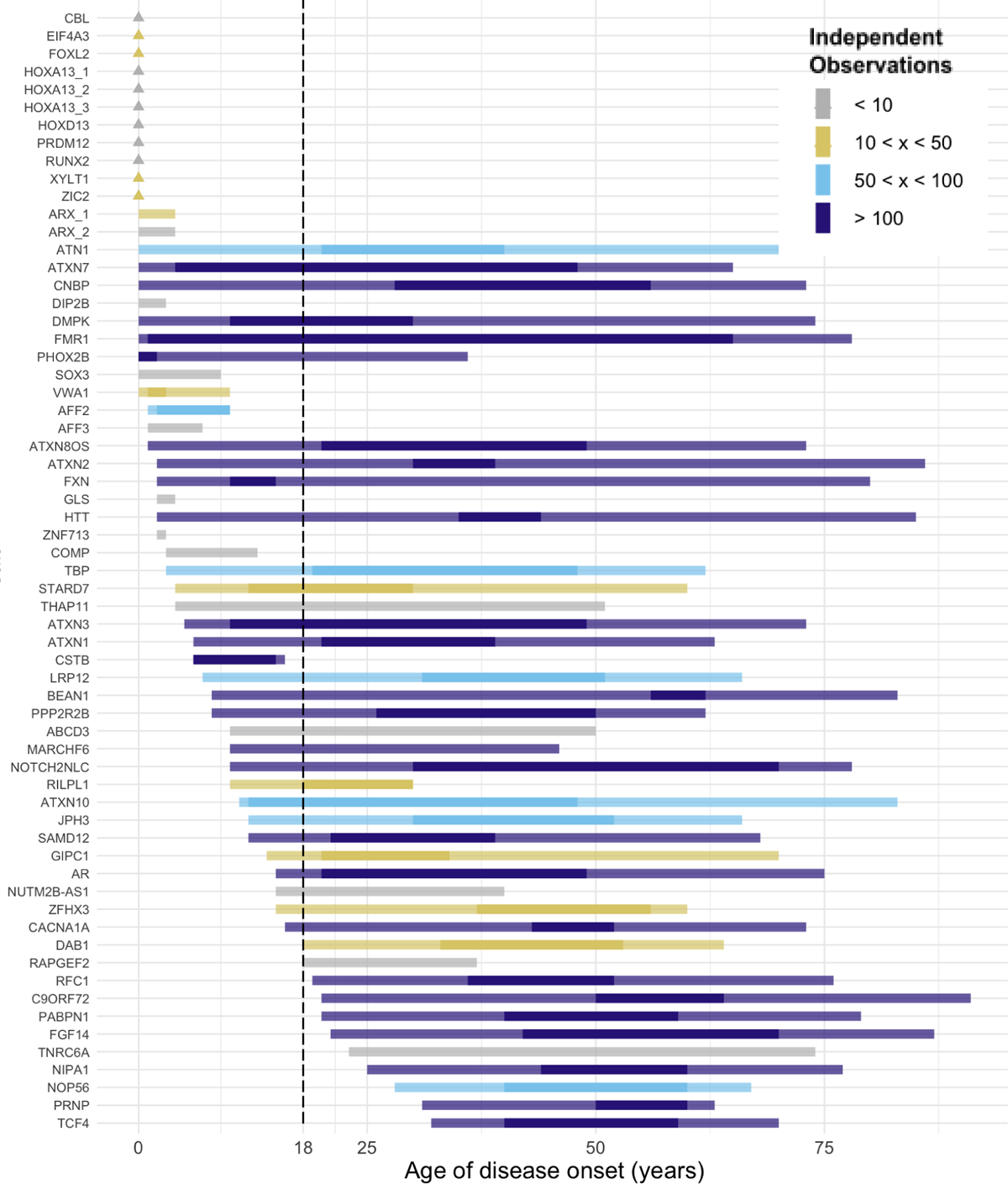
Ages of onset for TR disease range across the lifespan, with the majority of loci having possible pediatric onset. Triangles indicate congenital conditions occurring at birth. Lighter bars connect maximum and minimum reported ages, while opaque lines indicate typical intervals for age of onset, where greater than ten independent observations are available. Loci with less than two independent observations were removed, as was *POLG (*see **Methods**).

### Sequence motif complexity is essential to variant interpretation

STRchive annotates motifs detected at each locus by disease-relevant classification: benign, pathogenic, or uncertain significance. For most loci (59/68), the repeat motif in the reference genome (i.e., “reference motif”) is the pathogenic motif, and pathogenicity is determined exclusively by allele size. In the remaining nine loci, the observed motifs differ in pathogenicity, and specific patterns in the expansion may be necessary to cause disease.

Some motifs might expand without introducing pathogenicity, while others introduce pathogenicity at lower thresholds.^21,35^ For this reason, we document the locus structure or repeating sequence pertinent to disease for each locus. Although motif consideration is essential in variant interpretation, the biological consequence of motifs is still unknown in the majority of cases. Such subtleties may be overlooked in clinical evaluation and can introduce challenges in PCR-based assays.

### STRchive contextualizes gnomAD population data when assessing TR disease loci

A rational approach to elucidating the details of TR loci, such as motif significance or allelic frequency is by investigating population-level TR data.^17^ Empowering such an analysis, gnomAD v3.1.3 recently added allele size estimates at 60 disease-associated TR loci from more than 18,000 individuals using ExpansionHunter; this data is a subset of gnomAD individuals where whole-genome sequencing data was available for TR variant calling. As most TR data are derived from case studies or small cohorts of affected individuals, this database is an invaluable step forward to elucidate locus-specific variation in the general populace. At the same time, each locus presents unique bioinformatic and biological contexts which are necessary to understand when performing variant-, locus-, and phenotype-based analyses.

We leverage comprehensive, locus-specific information from STRchive to assess the gnomAD genotypes, which include motif and allele size estimates. We estimate the fraction of gnomAD population with pathogenic genotypes (PG) and with carrier status, taking inheritance patterns into account. Only calls where the sequenced motif matched a pathogenic motif are considered pathogenic. We exclude loci genotyped with “CGN”, as these were shown to have inflated allele estimates likely due to non-specificity of the “N” and proximity to other repetitive sequences (**Supplementary Methods**). Given the intrinsic complexity of TR diseases, some simplification was used. Expansion is typically considered necessary for TR pathogenicity. However, loci such as *VWA1* have suggestive evidence of pathogenicity with any deviation from a constrained norm, which includes either expansions or contractions.^36,37^ As there is limited evidence for the likelihood of pathogenicity with contractions, the role of modifier alleles, and other such biological circumstances, our analysis was restricted to allelic expansions with pathogenic motifs at non-“CGN” loci (**Methods**).

We identify 17 autosomal dominant loci with at least one expanded allele and three X-linked recessive loci with either one expanded allele in males (*DMD, AR*) or two expanded alleles in females (*DMD*) (Figure 4). Results are contrasted with general disease prevalence in the literature where available (citations available in STRchive documentation) and demonstrate cases of robust overlap (such as *TCF4, HTT,* and *ATN1)* and cases of separation (*DMD*, *ATXN8OS*, *PABPN1)* which in turn could imply reduced penetrance, delayed onset, or even questionable pathogenicity. Full calculated results are available in Supplementary Table 1.

**Figure 4:**
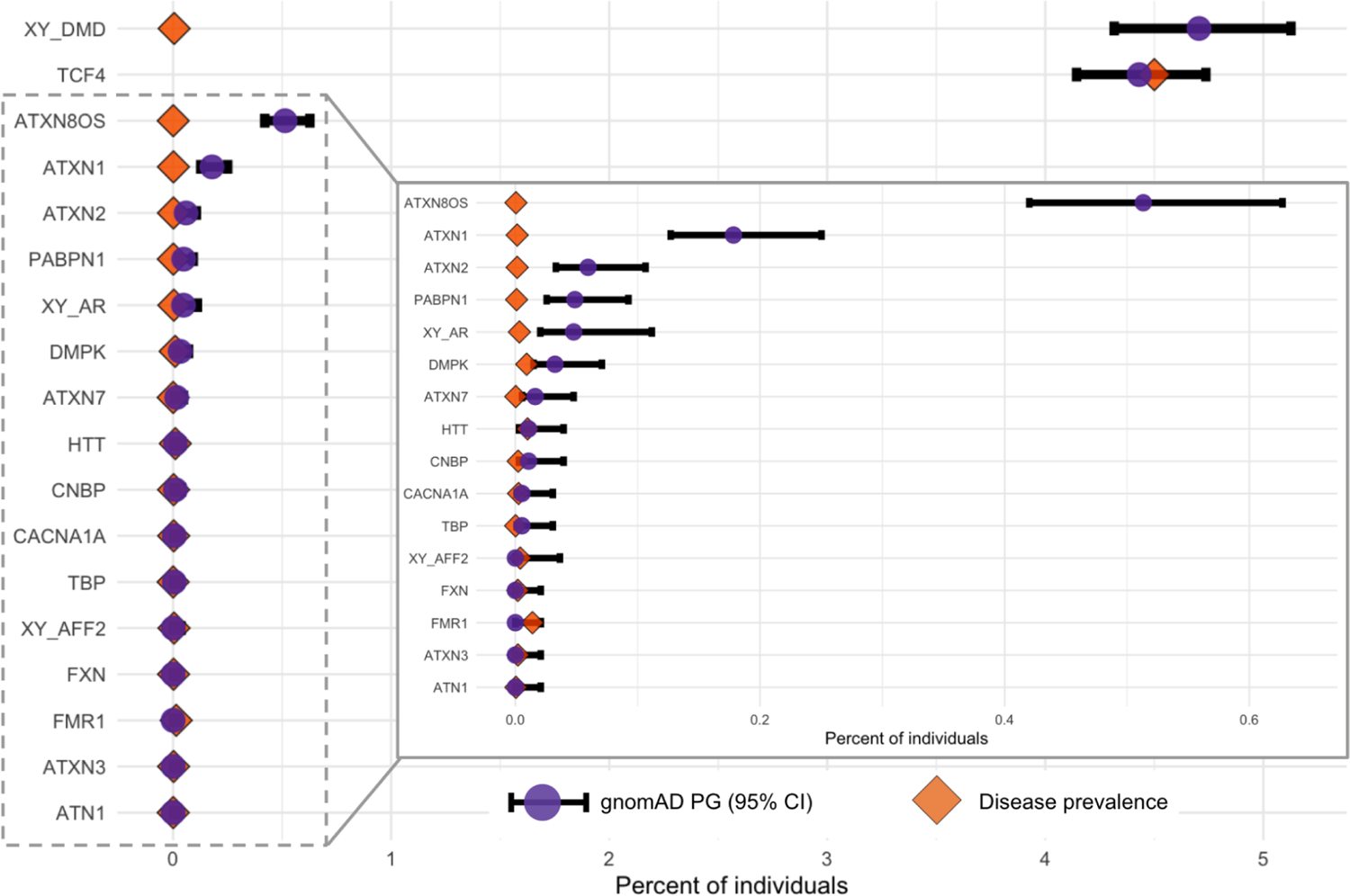
Pathogenic genotypes are found within the presumably unaffected gnomAD cohort, which correspond to and vary from known prevalence dependent on loci. Disease loci where PGs were found have the PG percentage (purple circle) within the gnomAD cohort shown, compared to disease prevalence ascertained by the literature (orange diamond). The PG percentage has a 95% binomial confidence interval calculated and plotted (black bar). Loci where prevalence is unknown are excluded. The inset plot’s x-axis is 0.0-0.64.

We now demonstrate the application of STRchive to the diagnostic process by discussing loci within the gnomAD dataset that exhibit unique aspects of TR variant interpretation, noting how these vignettes intersect with our variant interpretation guideline (Table 1). Our guideline and clinical vignettes reflect three overarching themes: evaluating allele(s), evaluating phenotype, and evaluating the locus.

**Table 1:**
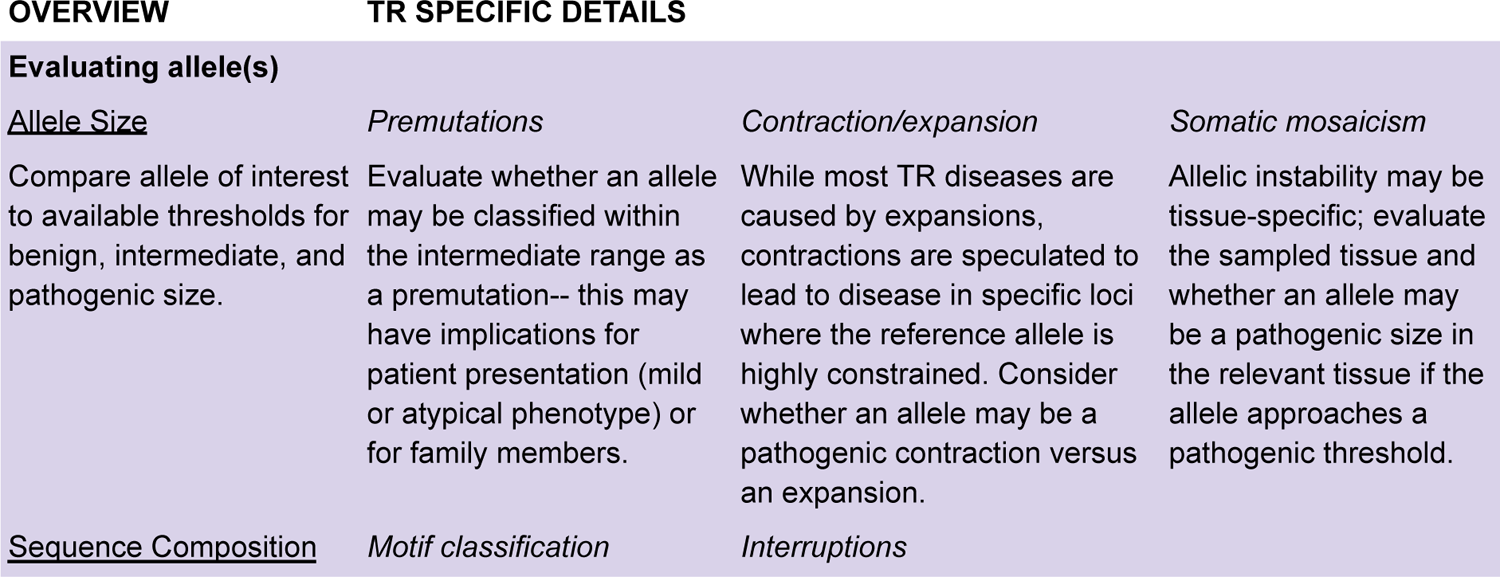
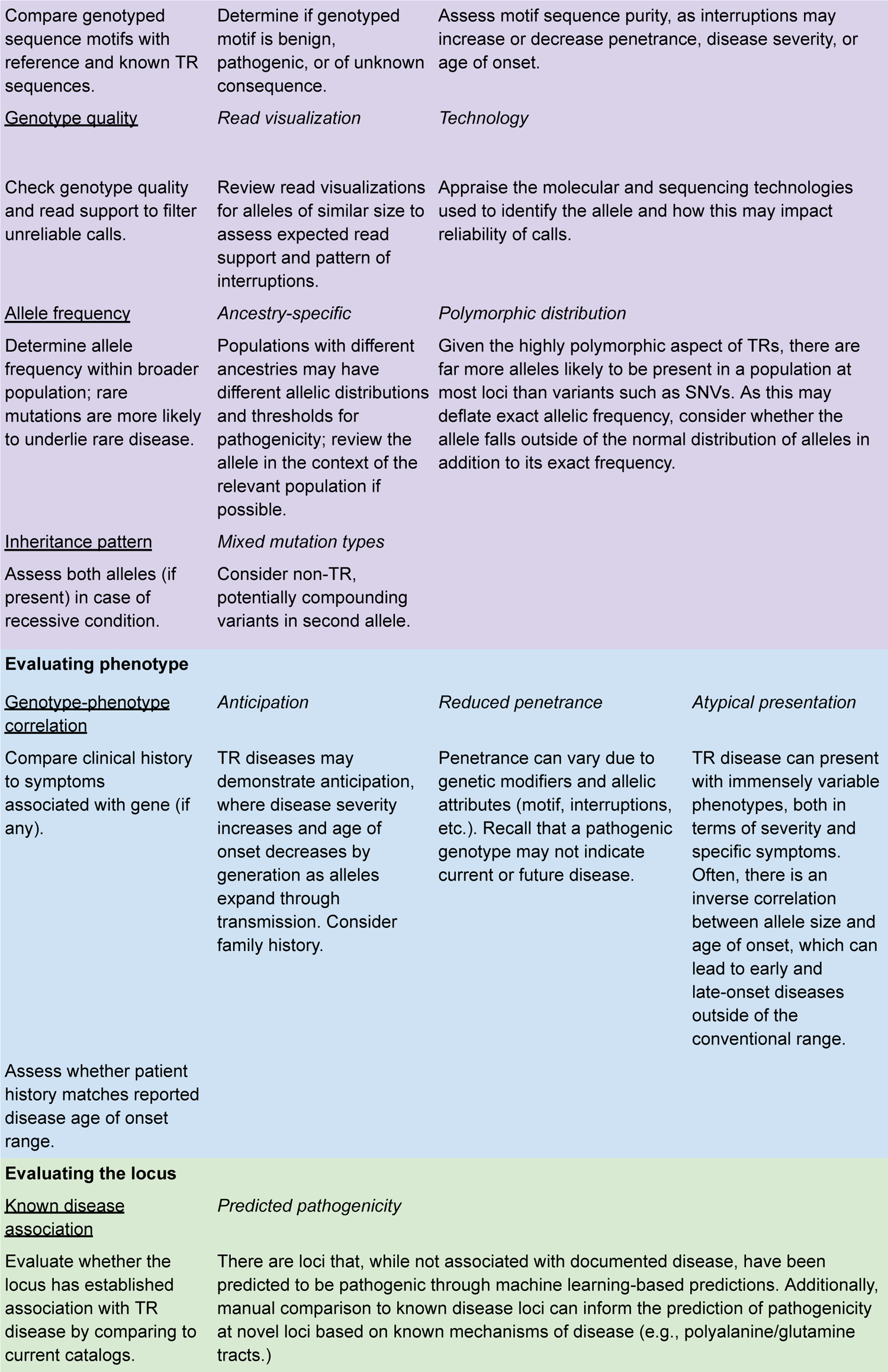
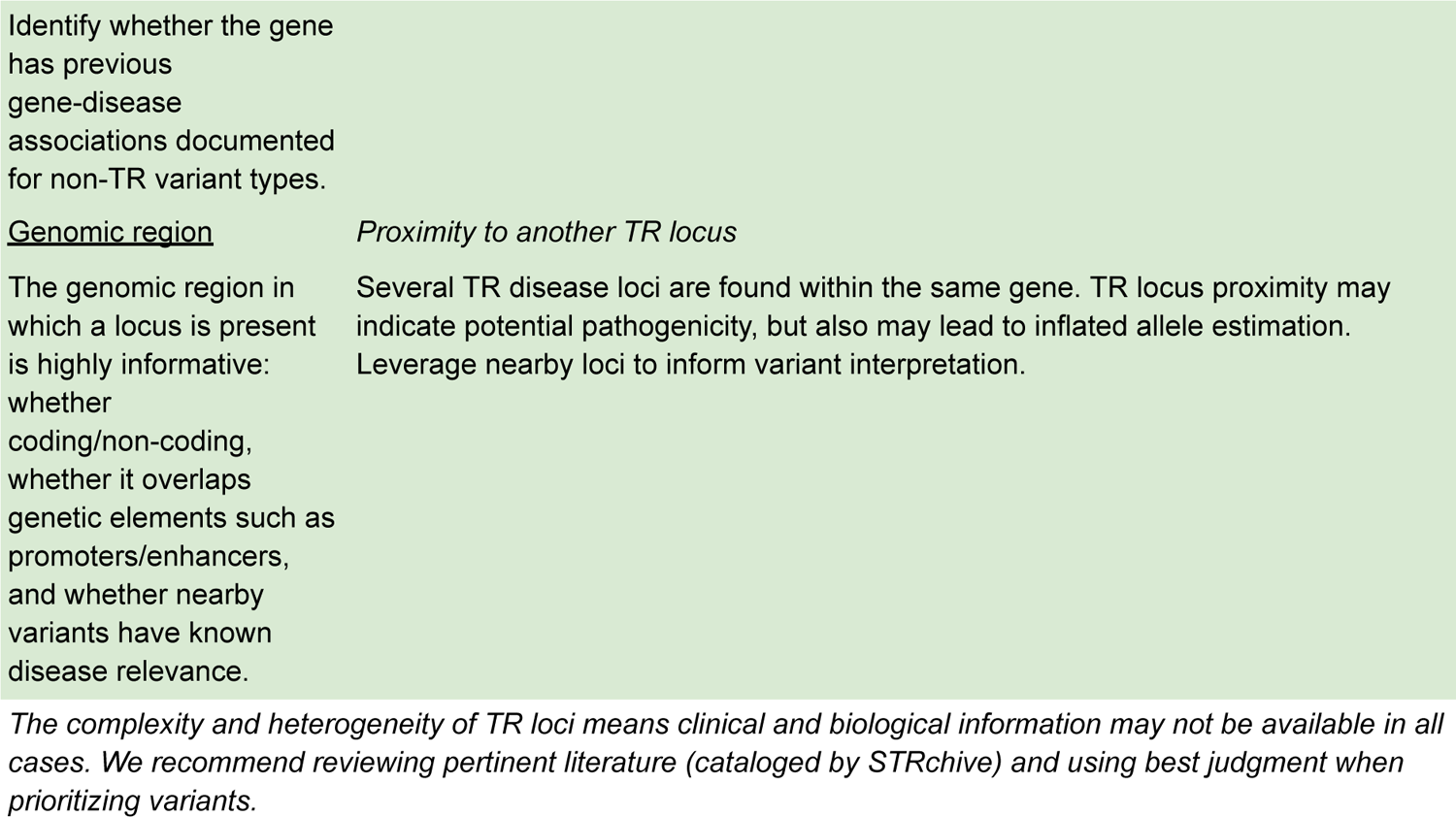
STRchive provides a blueprint to aid variant interpretation in a diagnostic workflow. A version of the blueprint that links current resources relevant to each point is available in Supplementary Table 3.

### Evaluating allele(s)

STRchive integrates literature and resources related to *allele frequency*, *inheritance patterns*, and methods of assessing *genotype quality*, in addition to carefully curated information related to **allele size** and **sequence composition**.

### **Allele size** can profoundly inform clinical expectations

TR disease loci are often evaluated in a binary fashion: if the allele exceeds a pathogenic threshold (or two alleles in a recessive condition), it is considered a pathogenic genotype. However, exact allele size is an essential consideration in interpretation, as age of onset and disease severity can be highly variable and correlated with repeat length (Figure 3). For example, while HD most typically presents in adults of three to four decades, sufficiently large expansions can cause disease onset in children as young as three years, while smaller pathogenic expansions may lead to disease in elderly individuals with mild symptoms.^38^ In gnomAD, 0.011% (95% confidence interval: 0.003–0.0394%, Supplementary Table 2) of individuals had at least one *HTT* allele exceeding 39 repeats, which closely matches the prevalence documented in the literature of 0.0106–0.0137%.^17,39^ The presence of PGs in the gnomAD cohort, even with conservative genotype estimates, may reflect the presence of these minimally expanded variants (mean of expanded alleles: 42 repeats) leading to patient ascertainment at a presymptomatic age.

While not ubiquitous, the relationship between allele size and clinical outcome is observed across many TR disease loci.^8^ Spinocerebellar ataxia 8 (SCA8) is caused by a CTG expansion and a corresponding, complementary CAG expansion in the overlapping *ATXN8OS* and *ATXN8* genes, respectively.^40^ The observed range of pathogenic alleles causing SCA8 is notably wide (71–1,300 repeats) and allele length is believed to influence disease penetrance, severity, and progression.^41,40,42^ The SCA8 PG percentage in gnomAD is the second highest frequency for autosomal dominant loci at 0.513% (∼1 in 200 individuals, 95% CI: 0.4201–0.627%), a frequency 1000-fold higher than the estimated literature prevalence for SCA8.^17^ This incongruity reinforces previous research that expanded alleles greatly outnumber disease cases due to reduced penetrance, with intermediate and pathogenic range expansions occurring 1 in 100–1200 chromosomes, depending on the population.^42^ As such, comparing the magnitude of an allele against the patient’s age and clinical history is highly informative in the diagnostic process for these and other loci. Referencing the clinical literature cataloged by STRchive can provide points of comparison to set expectations of phenotype.

### **Sequence composition** is an essential aspect of allele interpretation

At least 20 disease loci have shown clinically relevant changes in sequence composition, whether dispersed within a sequence as interruptions, alternating with the canonical motif, or entirely replacing the reference allele with an alternative motif.^43^ As such, STRchive documents motifs and records pertinent interruptions as they affect sequence composition, which in turn can impact patient phenotype. Within the gnomAD data set, exactly 15% of loci (9/60) had multiple motifs (2–20) genotyped beyond the reference (Figure 5). The *RFC1* locus underlying cerebellar ataxia, neuropathy, and vestibular areflexia syndrome (CANVAS) had 20 unique motifs identified with pathogenic motifs with relatively common frequency (**Supplementary** Figure 2).

**Figure 5:**
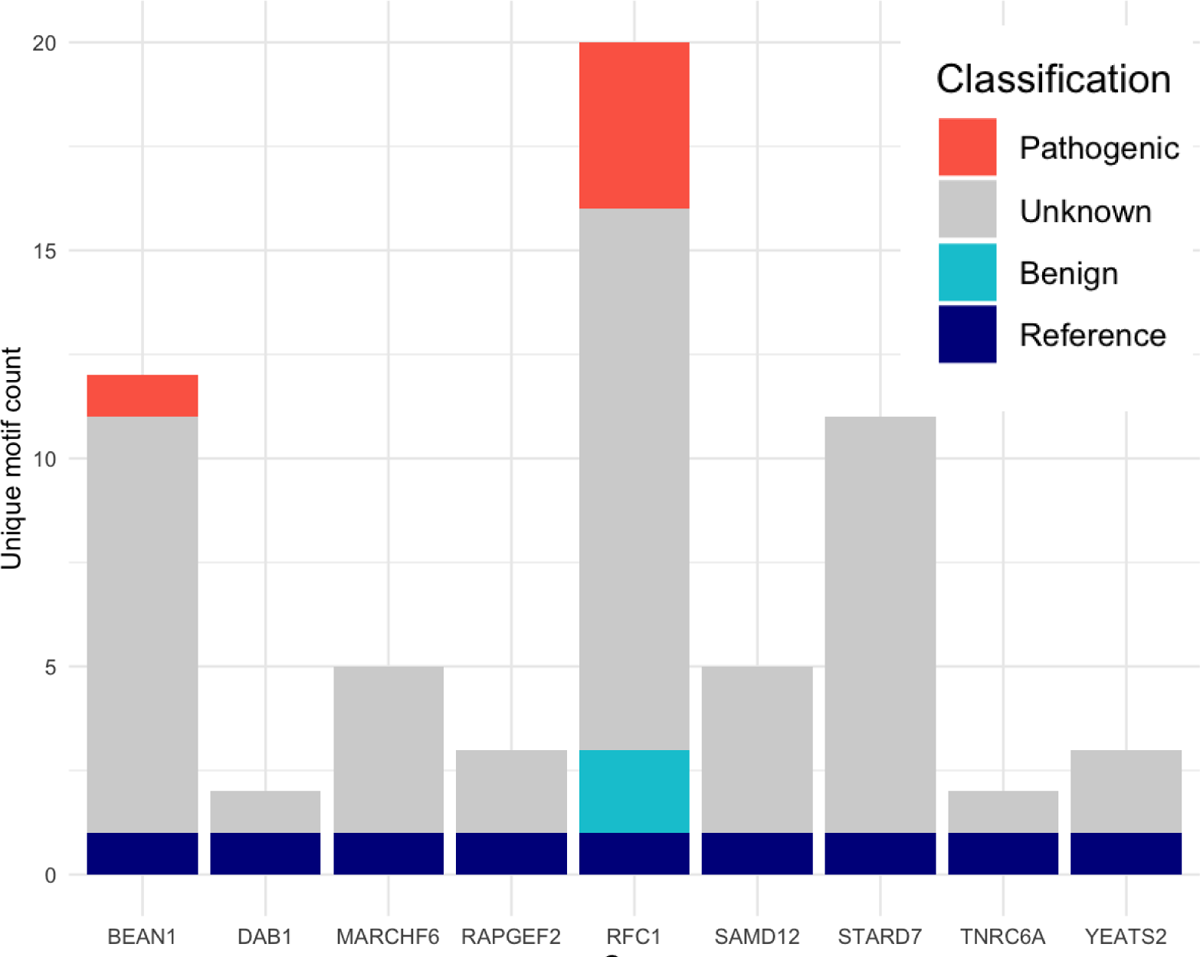
Nine gnomAD loci demonstrate motif heterogeneity, with two possessing pathogenic motifs captured in locus genotypes. Unique gnomAD motif counts where greater than one motif (the reference motif) is present, with STRchive motif classification applied.

The motif diversity at TR loci adds complexity to variant interpretation and is an ongoing area of development, as reflected in our data. Motif consequence is unknown in about 3/4ths of distinct motifs detected at these nine loci (47/63 unique motifs genotyped). Without knowing the association between a motif and a phenotype, or the threshold at which pathogenicity occurs for a specific motif, allelic consequence is challenging to determine.

Motif heterogeneity is common even within a smaller cohort: we identify unique motifs from 100 individuals from the Human Pangenome Reference Consortium (HPRC) genotyped by TRGT.^44^ Six gnomAD loci with multiple motifs also had multiple motifs in the HPRC data (*BEAN1, RAPGEF2, RFC1, SAMD12, STARD7, YEATS2;* **Supplementary** Figure 3). Four additional loci showed motif heterogeneity in the HPRC data (*FGF14, XYLT1*, *ZFHX3, C9orf72*), with none of the non-reference motifs of these four loci having documented classification in STRchive (**Supplementary** Figure 4). These findings highlight the importance of ongoing motif documentation within STRchive as information becomes available about motifs’ phenotypic implications.

In addition to motifs, interruptions within a sequence can greatly impact phenotype. *ATXN8OS* interruptions are known to influence disease status and severity in SCA8.^40,45^ Specifically, interruptions within the CAG tract appear to increase penetrance and protein toxicity.^45^ As affected and unaffected individuals can have *ATXN8OS* expansions (as reinforced by our dataset),^42^ the SCA8 locus further exemplifies the need to consider sequence composition in variant interpretation. Sequence composition changes may complicate variant interpretation on a bioinformatic level by impacting detection performance and genotyping accuracy.^14,15^ Interruptions may inflate allelic estimate, and an expansion may be missed if the correct motif is not targeted during genotyping.^9^ By documenting sequence composition changes, STRchive endeavors to facilitate TR detection in addition to aiding diagnosis.

### **Allele frequency** within a population can inform expectations of pathogenicity

Although we do not evaluate the exact allelic frequency of TRs within a population given their polymorphic nature, we assess the frequency of PGs in a population presumed to be unaffected by TR disease. While gnomAD is among the largest TR cohorts to date, TR diseases are rare and each specific disease typically affects far fewer than one in 20,000 individuals. Thus, most disease loci with full penetrance would not be expected to have PGs in this cohort of ∼18.5K individuals. Of the four disease loci where a PG is feasible by prevalence alone (estimated ≥1 in 18,500: *DMPK*, *HTT*, *FMR1*, *TCF4*), all but *DMPK* had a PG confidence interval spanning the documented literature prevalence. This highlights the necessity of considering allele frequency specifically, rather than solely disease prevalence: our *DMPK* findings (0.0324%, 95% CI: 0.0149–0.0707%) are comparable to one study’s frequency of *DMPK* repeat expansions taken from more than 50,000 newborn screenings (0.0476%, 0.0286–0.0667%).^46^ This suggests that *DMPK* expansions are present in the general population even at birth, and may pose as incidental or secondary findings. While genotyping inaccuracy in particularly large alleles could potentially lead to size underestimation, all *DMPK* PGs in the gnomAD cohort are within the “mild” expansion range of the disease, which can present as late as age 70.^47^

The gnomAD *DMPK* data also matches prevalence estimates ascertained within specific populations of elevated prevalence (such as Iceland), which may indicate population specificity which in turn can result in different allele frequencies.^47,48^ Allelic frequencies should be considered in the context of patient ancestry, which may impact the distribution of TR variant sizes. However, prevalence rates and allele frequency estimates are unavailable for many TR disease loci given heterogeneous clinical presentations, variable population ancestries, and technical limitations.^22^ Rarer TR diseases likely require a larger population cohort for sufficiently granular resolution establishing allelic frequency as well as more certainty about genotype accuracy to meaningfully compare to prevalence.

Disparities between large cohort PGs and clinically based disease prevalence estimates have been noted previously. In a study leveraging TOPMed and the 100,000 Genomes Project (100kGP) to genotype STR disease loci across ∼50k individuals, Tucci et al. estimated that TR diseases likely affect up to three times more individuals than currently recognized clinically.^22^ Of the thirteen loci surveyed by Tucci et al. also in the gnomAD dataset, nine had PG estimates concordant with our data, defined as a cohort estimate within or within 0.001% of the gnomAD 95% confidence interval (**Supplementary** Figure 5**, Supplementary Table 4**). Three loci were concordant once the distinct pathogenic thresholds used by Tucci et al. were applied to the gnomAD data. Only one locus remained discordant: *FXN*, known to have ancestry-specific disease prevalence.^72^ To resolve the ambiguities presented by the above discordances and associated research, STRchive will continue to record prevalence estimates and allelic frequencies as derived, which can be used in turn to evaluate the likelihood of a variant’s pathogenicity.

### Evaluating phenotype

STRchive catalogs extensive literature describing clinical cases and assorted **genotype-phenotypes**. Links to important clinical resources specific to TR diseases are provided within the website, as are comments on factors that may precede atypical clinical presentations (Supplementary Table 3). STRchive locus definitions redirect to specific locations within the UCSC Genome Browser, which itself shows overlapping gene phenotypes and can be overlaid with informative tracks.

### Informed **genotype-phenotype** comparisons can lead to candidate inclusion (or exclusion)

Carefully evaluating alleles of interest can inform expectations for phenotype, such as in HD when there is remarkably early- or late-onset of disease based on allele size. Similarly, awareness of changes in sequence composition can explain atypical presentations; for example, “CCG” interruptions within the “CTG” STR expansion in *DMPK* lead to unusual disease traits such as severe axial and proximal weakness, in addition to delayed onset of symptoms.^9^ Interruptions such as these may explain some of the presence of *DMPK* PGs in gnomAD exceeding disease prevalence. *Trans*-genetic elements may modify disease presentation, including non-TR mutations in related genes,^49^ and epigenetic factors like methylation can influence allele penetrance.^7,23^ There may be phenotypic considerations at a loci that extend beyond the allele to the overall disease. “Atypical” presentations may be the norm for loci with tremendous clinical heterogeneity: *NOTCH2NLC “*CGG” expansions are associated with neuronal intranuclear inclusion disease, Alzheimer’s disease, essential tremor, Parkinson’s disease, amyotrophic lateral sclerosis, and oculopharyngodistal myopathy.^50^ Additionally, some loci exhibit anticipation or a worsening of phenotype over generations, increasing the utility of family history. Lastly, reduced penetrance may lead to the complete absence of phenotype even when an expansion is observed. These considerations are complex, and we endeavor to provide robust resources through STRchive to distinguish between non-causative expansions versus expansions leading to atypical phenotypes, as well as flag loci with anticipation and reduced penetrance to inform diagnostic expectations.

Beyond specific symptom matching, evaluating the phenotype of a TR expansion can inform variant prioritization based on expectations of severity. The prevalence estimates documented by STRchive can underscore locus expectations: higher prevalence generally indicates a less deleterious disease. This trend was reflected in the gnomAD data: the highest percentage of PGs in an autosomal dominant condition and the second highest overall frequency in our dataset was 4.31% in *TCF4*, an STR locus causing Fuchs endothelial corneal dystrophy 3 (FECD3).^51^ FECD3 is estimated to affect 4% of the population older than 40 years, with a decades-long disease progression leading to reduced endothelial function and vision impairment. In contrast to many other TR diseases, corneal dystrophy is not expected to reduce lifespan or reproductive success. In fact, FECD3 was originally overlooked as a pathogenic expansion because neurodegeneration was the expected phenotype of an STR-associated disease, leading to the assumption that this variant was benign and unrelated to corneal dystrophy.^49^ Most patients with FECD3 show expanded alleles (68–76%), but penetrance is incomplete, as expanded alleles are also found in 3–6% of unaffected individuals. As such, the 4.31% PG percentage for *TCF4* in the gnomAD cohort is plausible.

Conversely to the late-onset FECD3, Duchenne muscular dystrophy (DMD) is a severe, progressive disease with motor symptoms typically by age 2–3. Most patients are wheelchair dependent after the first decade of life. One published report links an STR expansion to DMD, and the *DMD* locus is thus included in catalogs of TR diseases such as gnomAD. Given the early onset of DMD, it would be an unexpected causative variant in an adult patient. Similarly, we expect no *DMD* PGs in the gnomAD cohort, although females might be carriers of expanded alleles (≥59 repeats). The expected absence of *DMD* is furthered by its relatively rare prevalence: <1 per 10,000 in males and <1 per million in females.^52^

Instead, the *DMD* locus in males has our study’s highest PG percentage (4.705%, ∼1 per 20 males). A PG is identified in 0.089% of gnomAD females (∼1 per 1,000), and 8.198% of females are carriers of an expanded allele. Furthermore, the presence of expanded alleles across cohort sex is replicated in the long-read HPRC data. Two males (2/52, 3.85%) and two females (2/48, 4.17%) had PGs in the this dataset, and six females were carriers (12.5%). These data contrast dramatically with the disease prevalence of <1 per 10,000 in males and <1 per million in females.^52^

### Evaluating the locus

In addition to evaluating a specific variant, we can also leverate STRchive to evaluate whether a locus is truly disease relevant. We report the independent observations associated with each locus in addition to the number of PMIDs to show the general level of evidence for each disease (Figure 2). The well-studied *HTT* locus linked to Huntington’s disease has notably more publications than any other locus, with thousands of cases supporting its characterization. In contrast, more recently discovered loci such as *STARD7* or contested loci such as *DMD* have far fewer associated PMIDs and independent observations. By assessing a TR variant alongside its locus, diagnostic teams can prioritize and deprioritize putative variants as appropriate.

### Evidence of TR clinical relevance varies substantially by locus

Presented with a disease of early, severe symptoms juxtaposed with an insupportably high PG percentage (4.705/0.089% in gnomAD and 3.85/4.17% in the HPRC data in males and females, respectively), it is worth evaluating the validity of a causal role for the STR expansion at the *DMD* locus.^15^ The proposed PG percentages in the short- and long-read data become even more inconsistent with population prevalence estimates when considering the contribution of other variant classes within *DMD* gene as a whole to the overall disease burden. The majority (∼2/3rds) of causative variants underlying DMD are deletions of one or more exons, with the second greatest pathogenic contribution from partial duplications (∼10%), and then, other variant classes such as missense variants.^53^ We would expect STR expansions causing DMD to be far rarer than the general prevalence of DMD, given the commonness of other variant types. These STR expansions being far more common than the prevalence of all pathogenic DMD variants combined indicates that expansions at the *DMD* locus are unlikely to be pathogenic.

As such, it is necessary to interrogate the *DMD* TR locus and its proposed disease relevance. The primary non-experimental method to do so is literature review, which is facilitated by STRchive’s automated literature retrieval. *DMD* a highly repetitive gene, and cataloged literature discuss TRs as markers in linkage and carrier analysis. Nevertheless, only the single case report identifies a “dynamic” expansion of 59–82 repeats through three generations of a pedigree segregating DMD.^54^ The impact of this variant on the disease phenotype is speculated without mechanistic validation. No additional studies support the contribution of STR expansion on a DMD phenotype, even when assessing over a thousand individuals with hundreds of heterogeneous variants.^55,56^ In fact, a study genotyping long-read data from 878 individuals within the 1000 Genomes Project found 28 males (6.53%, 28/429) and four females (0.90%, 4/446) with theoretically pathogenic genotypes, as well as 21 female carriers (4.71%) (**Supplementary Methods**).^57^ We thus present an additional cohort analysis to refute *DMD* as an STR disease loci, the largest such study to date. Our evidence for refuting the *DMD* STR locus’ role in disease underscores the need for a responsive and dynamic database of STR disease loci that can integrate up-to-date information to ensure reliability.

Although the singular report of *DMD*’s TR association is disparate from established disease loci such as *HTT* and *C9orf72* (Figure 2), there are additional loci with limited literature such as *ZIC2*, *AFF3*, and *ZNF713*. Furthermore, novel loci will continue to be discovered and require interrogation despite an absence of comparative data. Innovative strategies may be necessary to evaluate pathogenicity, such as assessing genomic region (e.g., coding versus non-coding, overlap with genetic elements) and gene association with disease for nearby non-TR variants. Pathogenicity may also be predicted by tools such as REXprt ^58^. Ultimately, clinical teams must exercise best judgment and leverage available literature and databases when prioritizing likely TR variants. STRchive consolidates these resources to expedite locus and variant analysis, and will mature alongside the TR field.

### Availability and potential impact of STRchive

We provide query code (AutomatedLiteratureRetrieval.R in the STRchive_manuscript GitHub) and up-to-date literature directories for the convenience and benefit of STRchive users (http://strchive.org/). We have distilled pertinent information into comprehensive CSV and JSON files and a website-comprehensive table for easy user access. These catalogs will consistently evolve to capture updated loci and facilitate clinical and research endeavors. A version of our workflow has been integrated into the Utah NeoSeq project, a collaboration between the Utah Center for Genetic Discovery and ARUP Laboratories to diagnose Neonatal Intensive Care Unit patients, as well as into the Undiagnosed Diseases Network, a project funded by the National Institutes of Health to identify genetic etiologies for long-term undiagnosed conditions.^59,60^

## Discussion

STRchive is a comprehensive yet digestible resource of TR Mendelian disease loci. Given its infrastructure within GitHub, STRchive is poised for ongoing revision. Our database can quickly and easily incorporate vetted community contributions outside of regular maintenance to avoid the frustrations of “abandonware.”^14^ Even so, STRchive is a manually curated database of a rapidly evolving field. Although information is cited and cross-referenced across resources and by multiple experts, these data are snapshots of TR biology and clinical understandings, subject to clarification and evolution as research progresses. We are not exempt from the abounding complexities of TR genetic variation; users should check underlying evidence linked in STRchive and present in our collected literature. Concerning the aggregate cohort of gnomAD, we lack granular data such as age and PCR status for the majority of samples that could otherwise discretize our analysis of presumably non-penetrant expanded alleles. We also lack genotype data from some STRchive loci not present in gnomAD, precluding PG analysis at these loci.

### Capturing complexity for diagnostic empowerment

Almost half of STRchive 1.0.0 loci are exonic trinucleotide repeats, which may reflect a tendency in locus identification toward coding regions with comparable mechanisms to known diseases.^24,61^ However, as molecular and computational techniques develop, disease loci of greater unorthodoxy are likely to be discovered. In fact, the TR disease loci that have evaded discovery so far are likely to present with increased biological complexity, such as having multiple motifs, interruptions, allele size far exceeding the read length, occurrence at novel repeat loci, and complex locus structures.^33^ This shift is exemplified by recent discoveries such as the *RFC1* STR expansions causing CANVAS, which have multiple pathogenic motifs.^33^ RExPRT identified ∼30,000 TR loci in the genome as candidates for pathogenicity,^58^ suggesting that there are numerous additional disease loci and associated attributes to discover and integrate into STRchive.

TR pathogenic variants are proposed to explain some of the missing heritability in rare disease,^17,62^ in part because STRs have mutation rates that are orders of magnitude higher than any other variant class.^4,63^ Additionally, up to 70% of individuals with neurological conditions remain genetically undiagnosed,^8^ and TR disease loci are frequent causes of neuromuscular and neurodegenerative diseases. By improving the detection and interpretation of TR variants, clinical teams have the potential to provide informative diagnoses.^9^ STRchive offers expansive catalogs for multiple reference alignments designed to maximize variant capture. As new pathogenic loci are discovered (and documented within STRchive), their inclusion in rare disease workflows may lead to narrowed diagnostic gaps, clinically actionable outcomes, and shortened diagnostic odysseys.^15^ We anticipate that centralizing information within STRchive may improve the standardization of pathogenic thresholds across clinical laboratories, which, in turn, facilitates more efficient diagnostic processes.

Furthermore, we offer a diagnostic blueprint to guide clinical teams through evaluating allele(s) and prioritization of genotypes for further consideration (Table 1). Validation methods are frequently used to confirm TR expansions,^13,64,65^ and intentional evaluation as outlined can prioritize variants warranting resource-intensive followup. We provide evidence to endorse TR inclusion in instances where they are often diagnostically excluded, such as in pediatric workflows due to concerns over secondary findings.

Specifically, studies often presume that TR diseases are high penetrance conditions, with adult onset and limited actionability. This has likely led to systematic underdiagnosis of TR diseases in children and young adults. However, TRs are a common and potentially disproportionate cause of phenotypes frequently found in pediatric disease, such as ataxia.^66,67^ Our data also indicate that the majority of TR diseases can have pediatric onset (Figure 3). With regard to actionability, some TR conditions have treatments in early stages of development that may benefit patients, and diagnosis may be useful for family planning.^68–70^ Lastly, ending the diagnostic odyssey and incorrect diagnoses is often of intrinsic value to patients. As such, testing of relevant TR loci should be incorporated where clinical symptoms warrant further interrogation.

### Inferences made possible through cohort data

We found PG percentages to be broadly higher than disease prevalences estimated for the general populace (Supplementary Table 2). There are multiple possibilities for this variation, both biological and technical. The documented pathogenic threshold may be inaccurately defined, or disease penetrance may be lower when alleles are only slightly above the threshold. Prevalence might vary by ancestry and gnomAD subpopulations could differ from general estimates; this mismatch is conceivable for *PABPN1,* which has prevalence estimates ranging from 1 in 100,000 to 1 in 600 dependent on the population surveyed.^71^ Modifier alleles or changes in sequence composition may lead to reduced penetrance or delayed disease onset.^17^ Finally, despite efforts to call all genotypes accurately, certain loci may be subject to increased error rates that require long-read sequencing or higher read coverage to resolve.

However, the concordance between PG estimates across the TOPMed, 100kGP, and gnomAD cohorts suggests these allelic frequencies are generally accurate and raises several considerations. *Firstly,* it exemplifies how pathogenicity thresholds for TR disease loci remain subject to ongoing investigation and debate while profoundly impacting results.^9^ Additional large-scale studies of diverse ancestries are necessary to fully characterize benign, intermediate, and pathogenic allelic ranges. *Secondly,* our work and that of Tucci et al. suggest that allele size alone may be insufficient to diagnose TR disease, as even expansions that are rare by allelic frequency are found in healthy controls.^58^ Population-scale characterization of expanded alleles at loci believed to be completely penetrant has revealed PGs in unaffected individuals, and again, further characterization is necessary.^14^ *Lastly,* the *FXN* result hints at the population-specific components of TR disease. While most TR loci expansions are observed across ancestries,^22^ TRs are observed to vary in frequency and length distributions across ancestral groups.^15^ Inconsistencies in pathogenic thresholds may partly be due to population-specific allele distributions and disease penetrance.^17^ While most population-scale studies to date have either focused on European ancestry cohorts or been limited by sequencing depth,^23^ STRchive is positioned to incorporate updates as the above considerations are resolved.

### The future of TR disease loci

The pace of TR discovery and characterization is likely to continue accelerating as sequencing and bioinformatic techniques further evolve.^49^ There are several immediate opportunities for innovation. TRs are found across the genome in low-complexity regions such as centromeres and telomeres, which are difficult to interrogate with short-read sequencing.^41^ Additionally, while long-read sequencing resolves the issue of expansions exceeding read lengths, it introduces new problems such as stutter^14,65^ and remains prohibitively expensive. In parallel with the evolution of molecular and computational techniques, studies evaluating control and disease cases to characterize human variation will elucidate known and novel loci alike. There may be opportunities to directly compare pathogenic and non-pathogenic cases in large population databases of diverse ancestries, such as All of Us.^73,74^ Additional features of repeat sequences, such as methylation and mosaicism, may be assayed as made possible by new technologies.^13^ Although most studies to date have been largely observational, it is conceivable that therapeutics development will follow the increased characterization of disease loci, particularly as pathogenic mechanisms become better understood.^7^ As a comprehensive and dynamic resource, STRchive is positioned to support current and future initiatives addressing TR disease, from empowering resolution to long-standing diagnostic odysseys to guiding projects currently in their infancy.

## Methods

### STRchive manual curation

STRchive manual curation began with TR-specific reviews^1,7^ in conjunction with existing genomic databases such as gnomAD, OMIM, and STRipy. We supplemented the result database with preprints and publications gleaned from manual curation (such as through Google Scholar and PubCrawler alerts), input from clinical and research collaborators, and presentations at publicized genetics conferences. Loci were compared to the Tandem Repeats Finder track in the UCSC Genome Browser to standardize locus definitions. STRchive locus definitions are generally comparable to those used by gnomAD with a few exceptions driven by capturing the Tandem REpeats Finder track (manuscript script CatalogDifferences.ipynb, **Supplementary Methods**). These exceptions are explicitly chosen to improve sensitivity when overlapping output from various methods, for example, allowing an imperfect repeat within the sequence when appropriate. While gnomAD locus definitions are calibrated to optimize ExpansionHunter genotyping accuracy, STRchive locus definitions endeavor for greater universality in application and broader allelic capture, which sometimes increases reference width. TRGT also functions at higher accuracy with wider locus definitions, as genotyping accuracy is reduced when the flanking sequence contains additional repeat variation. We provide TRGT-compatible genotyping input files within the STRchive database.

### Automated literature retrieval and STRchive additional curation

Literature was retrieved on April 30, 2024 by searching for genes and gene synonyms acquired through biomaRt in conjunction with tandem repeat-related search terms through the R library easyPubMed—explanation of query refinement and modification and assessment of earliest PubMed publication are available in Supplementary Methods.

Queried PMIDs were leveraged in addition to OMIM, GeneReviews, and Orphanet to establish ranges in age of onset (including full and typical), detected motifs with clinical classification, prevalence estimates as available, and number of independent observations. All data incorporated into STRchive and related analyses were restricted to clinical cases explicitly linked to TR expansion. Pathologies sharing an OMIM entry but not exclusive to TR expansion (such as glutaminase deficiency or Duchenne Muscular Dystrophy) were reviewed to include TR-specific clinical cases. When literature was unavailable through query (for example, case reports published before indexing or restricted by language/terminology retrieval), publications were independently retrieved and assessed through interlibrary loan. Specific citations underpinning disease prevalence estimates and ranges in age of onset are included in related STRchive text fields in the full database. Disease prevalences in STRchive are averaged to a singular value when ranges are presented without a consensus prevalence estimate.

Disease loci with <2 independent observations (*DMD, ZIC3, TNR6CA, YEATS2,* and *TBX1* as of April 15, 2024) were removed from Figure 1B and Figure 3, given a lack of literature consensus to support establishing a reference for these loci. Additionally, *POLG* was removed, given the presence of expansions commonly in control/healthy individuals.^36^

### Calculating and comparing PGs

We used the genotypes generated in gnomAD by Expansion Hunter at the intersecting STRchive loci to estimate inferred pathogenic genotypes (PGs) based on pathogenic thresholds. For the analyses, the inheritance pattern for *ATXN2* and *PABPN1* was assumed to be autosomal dominant (AD), even though autosomal recessive cases have been seen in certain contexts. All motifs were normalized (nucleotides arranged in alphabetical order) to facilitate motif matching, as genotypes were required to be called with known pathogenic motifs to be considered potentially pathogenic. Loci with the genotyped motif “CNG” were excluded from calculations due to apparent inflation in allele estimates likely due to sequence non-specificity; the results underlying this exclusion are discussed in the Supplementary Methods.

The intersected gnomAD/STRchive dataset was subset by inheritance pattern (AD, autosomal recessive, X-linked dominant, and X-linked recessive) and calculated according to inheritance pattern. Dominant conditions required a single allele to exceed the pathogenic threshold (pathogenic_min) and a matched motif. In contrast, recessive conditions in individuals with two alleles required two inferred pathogenic alleles (exceeding pathogenic minimum with matched motifs) to have an inferred PG.

The number of PGs was calculated and converted to a percentage with the number of PGs as the numerator and the number of individuals genotyped at the locus as the denominator. A 95% binomial proportion confidence interval for the PG percentage was generated in R by using the number of individuals genotyped for a locus as the number of “trials” and the number of PGs as the number of “successes.”

In our estimates of PGs, we used the allele lower bound estimates for each allele because while there is broad concordance between the genotype and the lower bound estimate (allele estimates were identical in 97.02% of calls for allele 1 and 94.13% of calls for allele 2), Expansion Hunter tends to overestimate alleles when erring and we endeavored to be conservative in our estimates of pathogenicity.^11^ Average difference between allele 1 and the lower bound estimate is 0.22 repeat unit for all calls, and 7.40 (range 1–251, median 6) for the subset where allele 1 is not equal to lower bound estimates. For allele 2, the average distance was 0.42 repeat units for all calls and 7.14 for the subset of non-identical values (range 1–267, median 5). A full analysis script, including merging with STRchive disease prevalence estimates, is available at CalculatingPGsandConfidenceIntervals.R within the manuscript GitHub.

Comparison with Tucci et al. data was performed by comparing their reported PG percentages for intersecting loci to our data set’s PG percentage confidence intervals. Evaluation of gnomAD PGs when matching pathogenic thresholds to those used by Tucci et al. were performed by identical scripts as in our analysis, with the pathogenic minimum substituted for the new thresholds.

### Blueprint

The diagnostic blueprint was created to synthesize current workflows and considerations implemented through partnerships with the Undiagnosed Diseases Network and NeoSeq.

## Data Availability

STRchive is licensed under a Creative Commons Attribution 4.0 International License. STRchive is available at http://strchive.org/, with comprehensive data, metadata, and processing scripts available at https://github.com/dashnowlab/STRchive. All scripts for manuscript data analysis and figure generation are available at https://github.com/dashnowlab/STRchive_manuscript; publicly available data used for analyses is also hosted on this GitHub. gnomAD tandem repeat data, including allele frequency distributions, per-sample genotypes, and other sample metadata, can be explored online at https://gnomad.broadinstitute.org/short-tandem-repeats?dataset=gnomad_r3 and is also available for download on the gnomAD website under “v3 Downloads > Short Tandem Repeats”:https://gnomad.broadinstitute.org/downloads#v3-short-tandem-repeats The long-read data from the Human Pangenome Reference Consortium is available from SRA project PRJNA701308 or https://humanpangenome.org/data/.

## Acknowledgments

HD is supported by 5K99HG012796-02. LH is supported by 1F30CA284847-01. HR, BW, and GV were supported by NHGRI grant U01HG011755. LH thanks Thomas J. Nicholas for his instrumental feedback on the manuscript, and Jason Kunisaki for his input on several figures.

## References

1. Depienne, C. & Mandel, J.-L. 30 years of repeat expansion disorders: What have we learned and what are the remaining challenges? Am. J. Hum. Genet. 108, 764–785 (2021).

2. Willems, T. et al. Genome-wide profiling of heritable and de novo STR variations. Nat. Methods 14, 590–592 (2017).

3. Chaisson, M. J. P., Sulovari, A., Valdmanis, P. N., Miller, D. E. & Eichler, E. E. Advances in the discovery and analyses of human tandem repeats. Emerg. Top. Life Sci. ETLS20230074 (2023).

4. Gymrek, M. A genomic view of short tandem repeats. Curr. Opin. Genet. Dev. 44, 9–16 (2017).

5. Gemayel, R., Vinces, M. D., Legendre, M. & Verstrepen, K. J. Variable tandem repeats accelerate evolution of coding and regulatory sequences. Annu. Rev. Genet. 44, 445–477 (2010).

6. Fotsing, S. F. et al. The impact of short tandem repeat variation on gene expression. Nat. Genet. 51, 1652–1659 (2019).

7. Hannan, A. J. Tandem repeats mediating genetic plasticity in health and disease. Nat. Rev. Genet. 19, 286–298 (2018).

8. Ibañez, K. et al. Whole genome sequencing for the diagnosis of neurological repeat expansion disorders in the UK: a retrospective diagnostic accuracy and prospective clinical validation study. Lancet Neurol. 21, 234–245 (2022).

9. Chintalaphani, S. R., Pineda, S. S., Deveson, I. W. & Kumar, K. R. An update on the neurological short tandem repeat expansion disorders and the emergence of long-read sequencing diagnostics. Acta Neuropathol Commun 9, 98 (2021).

10. Sachenkova Lundström, O., et al. WebSTR: a population-wide database of short tandem repeat variation in humans. J. Mol. Biol. 168260 (2023).

11. Weisburd, B., Tiao, G. & Rehm, H. L. Insights from a genome-wide truth set of tandem repeat variation. bioRxiv 2023.05.05.539588 (2023) doi:10.1101/2023.05.05.539588.

12. Yu, A. C.-S. et al. A Targeted Gene Panel That Covers Coding, Non-coding and Short Tandem Repeat Regions Improves the Diagnosis of Patients With Neurodegenerative Diseases. Front. Neurosci. 13, 1324 (2019).

13. Dolzhenko, E. et al. Characterization and visualization of tandem repeats at genome scale. Nat. Biotechnol. 1–9 (2024).

14. Tanudisastro, H. A., Deveson, I. W., Dashnow, H. & MacArthur, D. G. Sequencing and characterizing short tandem repeats in the human genome. Nat. Rev. Genet. 1–16 (2024).

15. Bahlo, M. et al. Recent advances in the detection of repeat expansions with short-read next-generation sequencing. F1000Res. 7, (2018).

16. Marwaha, S., Knowles, J. W. & Ashley, E. A. A guide for the diagnosis of rare and undiagnosed disease: beyond the exome. Genome Med. 14, 23 (2022).

17. Tang, H. et al. Profiling of Short-Tandem-Repeat Disease Alleles in 12,632 Human Whole Genomes. Am. J. Hum. Genet. 101, 700–715 (2017).

18. Treangen, T. J. & Salzberg, S. L. Repetitive DNA and next-generation sequencing: computational challenges and solutions. Nat. Rev. Genet. 13, 36–46 (2011).

19. French, C. E. et al. Refinements and considerations for trio whole-genome sequence analysis when investigating Mendelian diseases presenting in early childhood. HGG Adv 3, 100113 (2022).

20. Liu, Q., Tong, Y. & Wang, K. Genome-wide detection of short tandem repeat expansions by long-read sequencing. BMC Bioinformatics 21, 542 (2020).

21. Fazal, S. et al. Large scale in silico characterization of repeat expansion variation in human genomes. Sci Data 7, 294 (2020).

22. Tucci, A. et al. Population frequency of Repeat expansions indicates increased disease prevalence estimates across different populations. (2023) doi:10.21203/rs.3.rs-3097805/v1.

23. Shi, Y. et al. Characterization of genome-wide STR variation in 6487 human genomes. Nat. Commun. 14, 2092 (2023).

24. Panoyan, M. A. & Wendt, F. R. The role of tandem repeat expansions in brain disorders. Emerg. Top. Life Sci. ETLS20230022 (2023).

25. Rehm, H. L. et al. ClinGen--the Clinical Genome Resource. N. Engl. J. Med. 372, 2235–2242 (2015).

26. Landrum, M. J. et al. ClinVar: public archive of relationships among sequence variation and human phenotype. Nucleic Acids Res. 42, D980–5 (2014).

27. Adam, M. P. et al. GeneReviews®. (University of Washington, Seattle, 2024).

28. Amberger, J. S., Bocchini, C. A., Scott, A. F. & Hamosh, A. OMIM.org: leveraging knowledge across phenotype-gene relationships. Nucleic Acids Res. 47, D1038–D1043 (2019).

29. Halman, A., Dolzhenko, E. & Oshlack, A. STRipy: A graphical application for enhanced genotyping of pathogenic short tandem repeats in sequencing data. Hum. Mutat. 43, 859–868 (2022).

30. Weisburd, B., VanNoy, G. & Watts, N. The Addition of Short Tandem Repeat Calls to gnomAD. https://gnomad.broadinstitute.org/news/2022-01-the-addition-of-short-tandem-repeat-calls-to-gnomad/.

31. Kent, W. J. et al. The human genome browser at UCSC. Genome Res. 12, 996–1006 (2002).

32. Benson, G. Tandem repeats finder: a program to analyze DNA sequences. Nucleic Acids Res. 27, 573–580 (1999).

33. Read, J. L., Davies, K. C., Thompson, G. C., Delatycki, M. B. & Lockhart, P. J. Challenges facing repeat expansion identification, characterisation, and the pathway to discovery. Emerg. Top. Life Sci. ETLS20230019 (2023).

34. Maiuri, T. et al. DNA Repair in Huntington’s Disease and Spinocerebellar Ataxias: Somatic Instability and Alternative Hypotheses. J. Huntingtons Dis. 10, 165–173 (2021).

35. Dominik, N. et al. Normal and pathogenic variation of RFC1 repeat expansions: implications for clinical diagnosis. Brain 146, 5060–5069 (2023).

36. Eerola, J. et al. POLG1 polyglutamine tract variants associated with Parkinson’s disease. Neurosci. Lett. 477, 1–5 (2010).

37. Pagnamenta, A. T. et al. An ancestral 10-bp repeat expansion in VWA1 causes recessive hereditary motor neuropathy. Brain 144, 584–600 (2021).

38. Milunsky, J. M., Maher, T. A., Loose, B. A., Darras, B. T. & Ito, M. XL PCR for the detection of large trinucleotide expansions in juvenile Huntington’s disease. Clin. Genet. 64, 70–73 (2003).

39. McColgan, P. & Tabrizi, S. J. Huntington’s disease: a clinical review. Eur. J. Neurol. 25, 24–34 (2018).

40. Entry - #608768 - SPINOCEREBELLAR ATAXIA 8; SCA8 - OMIM. https://omim.org/entry/608768.

41. Rajagopal, S., Donaldson, J., Flower, M., Hensman Moss, D. J. & Tabrizi, S. J. Genetic modifiers of repeat expansion disorders. Emerg. Top. Life Sci. ETLS20230015 (2023).

42. Cleary, J. D., Subramony, S. H. & Ranum, L. P. W. Spinocerebellar Ataxia Type 8. (University of Washington, Seattle, 2021).

43. Rajan-Babu, I.-S., Dolzhenko, E., Eberle, M. A. & Friedman, J. M. Sequence composition changes in short tandem repeats: heterogeneity, detection, mechanisms and clinical implications. Nat. Rev. Genet. (2024) doi:10.1038/s41576-024-00696-z.

44. Wang, T. et al. The Human Pangenome Project: a global resource to map genomic diversity. Nature 604, 437–446 (2022).

45. Perez, B. A., et al. CCG•CGG interruptions in high-penetrance SCA8 families increase RAN translation and protein toxicity. EMBO Mol. Med. 13, e14095 (2021).

46. Johnson, N. E. et al. Population-Based Prevalence of Myotonic Dystrophy Type 1 Using Genetic Analysis of Statewide Blood Screening Program. Neurology 96, e1045–e1053 (2021).

47. Bird, T. D. Myotonic Dystrophy Type 1. (University of Washington, Seattle, 2024).

48. Yum, K., Wang, E. T. & Kalsotra, A. Myotonic dystrophy: disease repeat range, penetrance, age of onset, and relationship between repeat size and phenotypes. Curr. Opin. Genet. Dev. 44, 30–37 (2017).

49. Gall-Duncan, T., Sato, N., Yuen, R. K. C. & Pearson, C. E. Advancing genomic technologies and clinical awareness accelerates discovery of disease-associated tandem repeat sequences. Genome Res. 32, 1–27 (2022).

50. Zhang, T., Bao, L. & Chen, H. Review of Phenotypic Heterogeneity of Neuronal Intranuclear Inclusion Disease and NOTCH2NLC-Related GGC Repeat Expansion Disorders. Neurol Genet 10, e200132 (2024).

51. Entry - #613267 - CORNEAL DYSTROPHY, FUCHS ENDOTHELIAL, 3; FECD3 - MIM. https://omim.org/entry/613267.

52. Duan, D., Goemans, N., Takeda, S. ’ichi, Mercuri, E. & Aartsma-Rus, A. Duchenne muscular dystrophy. Nat Rev Dis Primers 7, 13 (2021).

53. Entry - #310200 - MUSCULAR DYSTROPHY, DUCHENNE TYPE; DMD - OMIM. https://omim.org/entry/310200.

54. Kekou, K. et al. A dynamic trinucleotide repeat (TNR) expansion in the DMD gene. Mol. Cell. Probes 30, 254–260 (2016).

55. Viggiano, E. et al. Spectrum of Genetic Variants in the Dystrophin Gene: A Single Centre Retrospective Analysis of 750 Duchenne and Becker Patients from Southern Italy. Genes 14, (2023).

56. Kekou, K. et al. Retrospective analysis of persistent HyperCKemia with or without muscle weakness in a case series from Greece highlights vast DMD variant heterogeneity. Expert Rev. Mol. Diagn. 23, 999–1010 (2023).

57. De Coster, W. et al. Medically relevant tandem repeats in nanopore sequencing of control cohorts. medRxiv 2024.03.06.24303700 (2024) doi:10.1101/2024.03.06.24303700.

58. Fazal, S. et al. RExPRT: a machine learning tool to predict pathogenicity of tandem repeat loci. Genome Biol. 25, 39 (2024).

59. Reynolds, H. M., et al. Rapid genome sequencing identifies a novel de novo SNAP25 variant for neonatal congenital myasthenic syndrome. Cold Spring Harb Mol Case Stud 8, (2022).

60. Murdock, D. R., Rosenfeld, J. A. & Lee, B. What Has the Undiagnosed Diseases Network Taught Us About the Clinical Applications of Genomic Testing? Annu. Rev. Med. 73, 575–585 (2022).

61. Hernandez, R. & Facelli, J. C. Structure analysis of the proteins associated with polyA repeat expansion disorders. J. Biomol. Struct. Dyn. 40, 5556–5565 (2022).

62. Maroilley, T. & Tarailo-Graovac, M. Uncovering Missing Heritability in Rare Diseases. Genes 10, (2019).

63. Mitra, I. et al. Patterns of de novo tandem repeat mutations and their role in autism. Nature 589, 246–250 (2021).

64. Yoon, J. G. et al. Diagnostic uplift through the implementation of short tandem repeat analysis using exome sequencing. Eur. J. Hum. Genet. 1–4 (2024).

65. Mastrorosa, F. K., Miller, D. E. & Eichler, E. E. Applications of long-read sequencing to Mendelian genetics. Genome Med. 15, 42 (2023).

66. Rafehi, H., Bennett, M. F. & Bahlo, M. Detection and discovery of repeat expansions in ataxia enabled by next-generation sequencing: present and future. Emerg Top Life Sci 7, 349–359 (2023).

67. Pavone, P. et al. Ataxia in children: early recognition and clinical evaluation. Ital. J. Pediatr. 43, 6 (2017).

68. Didonna, A. & Opal, P. The promise and perils of HDAC inhibitors in neurodegeneration. Ann Clin Transl Neurol 2, 79–101 (2015).

69. Srinivasan, S. R., Melo de Gusmao, C., Korecka, J. A. & Khurana, V. Chapter 18 - Repeat expansion disorders∗. in Neurobiology of Brain Disorders (Second Edition) (eds. Zigmond, M. J., Wiley, C. A. & Chesselet, M.-F.) 293–312 (Academic Press, 2023).

70. Leavitt, B. R. Chapter 24 - Current clinical trials of new therapeutic agents for Huntington’s disease. in Huntington’s Disease (eds. Yang, X. W., Thompson, L. M. & Heiman, M.) 571–589 (Academic Press, 2024).

71. Trollet, C. et al. Oculopharyngeal Muscular Dystrophy. (University of Washington, Seattle, 2020).

72. Bidichandani, S. I. & Delatycki, M. B. Friedreich Ataxia. (University of Washington, Seattle, 2017).

73. Koch, L. Global genomic diversity for All of Us. Nat. Rev. Genet. (2024) doi:10.1038/s41576-024-00727-9.

74. Manigbas, C. A. et al. A phenome-wide association study of tandem repeat variation in 168,554 individuals from the UK Biobank. medRxiv (2024) doi:10.1101/2024.01.22.24301630.

