## Supplementary Methods for "STRchive: a dynamic resource detailing population-level and locus-specific insights at tandem repeat disease loci"

### Automated Literature Retrieval

The automated literature retrieval script is available within the manuscript GitHub in the scripts folder as AutomatedLiteratureRetrieval.R. biomaRt is used to find gene synonyms of STRchive gene list, with a subset of synonyms excluded after a manual review of literature because of their overlap with other terms and conditions conflating search queries and returning multiple cases of irrelevant literature. Specifically:

B37: Synonym for *ATN1*, returns results on HLA-B37
MHP: Synonym for *CACNA1A*, returns results on medical health professionals, mental health problems, mental health professionals, etc.
MED: Synonym for *COMP*, returns results from medical journals (abbreviated to Med)
DM: Synonym for *DMPK,* returns results for diabetes mellitus
DM1: Synonym for *DMPK,* returns results for diabetes mellitus type 1
FA: Synonym for *FXN*, returns results for fatty acid and Fanconi anemia
GAC: Synonym for *GLS*, returns results for gastric adenocarcinoma
SPD: Synonym for *HOXD13*, returns results for spermidine, surfactant protein D, soluble programmed cell death protein 1 (sPD-1)
PRP: Synonym for *PRNP*, returns results for platelet-rich plasma
A1: Synonym for *RFC1*, returns results for HA1c, Apolipoprotein A1, A1 type fracture, etc.
CCD: Synonym for *RUNX2*, returns results for chronic coronary disease
PHP: Synonym for *SOX3*, returns results for palindrome-embedded hairpin structure
VCF: Synonym for *TBX1*, returns results for vertebral compression fracture and VCF file type

Similarly, *BMD* is substituted in the final query as “Becker muscular dystrophy” to avoid results related to bone mineral density.

The base function for getting PubMed results formulated the queries as follows:

```
query <- paste0('("repeat expansion"[Title/Abstract] OR "tandem repeat"[Title/Abstract] OR "repeat expansions"[Title/Abstract] OR "tandem repeats"[Title/Abstract] OR "repeat sequence"[Title/Abstract] OR "repeat sequences"[Title/Abstract] OR "repeat length"[Title/Abstract] OR "repeat lengths"[Title/Abstract] OR "expansion"[Title] OR "expansions"[Title] OR "repeats"[Title]) AND (', paste(joined_terms, collapse = " OR "),') AND "English"[Language] AND ("disease"[Title/Abstract] OR "disorder"[Title/Abstract] OR "diseases"[Title/Abstract] OR "disorders"[Title/Abstract] OR "syndrome"[Title/Abstract] OR "syndromes"[Title/Abstract] OR "patient"[Title/Abstract] OR "patients"[Title/Abstract] OR "proband"[Title/Abstract] OR "probands"[Title/Abstract]) AND ("journal article"[Publication Type] OR "letter"[Publication Type] or "Case Reports"[Publication Type]) NOT "review"[Publication Type]')
```

The query fundamentally contains four sections.

First, terms that are specific to tandem repeats: ("repeat expansion"[Title/Abstract] OR "tandem repeat"[Title/Abstract] OR "repeat expansions"[Title/Abstract] OR "tandem repeats"[Title/Abstract] OR "repeat sequence"[Title/Abstract] OR "repeat sequences"[Title/Abstract] OR "repeat length"[Title/Abstract] OR "repeat lengths"[Title/Abstract] OR "expansion"[Title] OR "expansions"[Title] OR "repeats"[Title])

Second, “joined terms” include genes and gene synonyms, grouped by overarching gene name defined as the HGNC symbol.

Third, clinical terms: ("disease"[Title/Abstract] OR "disorder"[Title/Abstract] OR "diseases"[Title/Abstract] OR "disorders"[Title/Abstract] OR "syndrome"[Title/Abstract] OR "syndromes"[Title/Abstract] OR "patient"[Title/Abstract] OR "patients"[Title/Abstract] OR "proband"[Title/Abstract] OR "probands"[Title/Abstract])

Fourth, terms to specify the type of returned articles: ("journal article"[Publication Type] OR "letter"[Publication Type] or "Case Reports"[Publication Type]) NOT "review"[Publication Type]')

Terms must be in the title and abstract of a publication to ensure topical publications are returned. Articles are also required to be in English, given the language proficiency of the STRchive team.

The earliest available publication indexed by PubMed was found by sorting output through the earliest publications and determining the PubMed ID (PMID) of the earliest article specific to the tandem repeat disease loci identified by STRchive. The earliest article was found for 57 out of 63 disease loci within our automatically retrieved PubMed results. For three loci (*SOX3*, *TBX1*, and *PRDMD12)*, the earliest PMIDs were not retrieved in our search because of a terminology mismatch that could not be corrected without dramatically inflating the number of unrelated publications in our results. For example, changing title-only terms such as “repeats” and “expansions” to also be queried in the abstract doubled the resultant PMIDs (04/04/2024), and adding polyglutamine and polyalanine to repeat expansion terms increased the PMID count by approximately a third overall. Two loci did not have their earliest PMIDs returned because of an atypical publication type (*CNBP*, *ATXN1*), and one article was not returned because its abstract was not indexed in PubMed (PPP2R2B). These articles were manually added to the publication data frame for visualization purposes.

### CNG Exclusion

We endeavored to restrict our PG analysis to high-confidence loci so that our conclusions could be robust. We conducted quality control analyses investigating the variation between genotypes when accounting for PCR status, incorporation of off-target regions, attribution to a public dataset, and motif. We found consistent evidence that loci genotyped with a “CNG” were error-prone. We determined that these eighteen loci (including three regions within *HOXA13* and two within *ARX*) should be excluded from this manuscript’s PG analysis.

Firstly, there appeared to be a genotyping error (shown in one locus for allele 2 but present across loci and alleles) where the allele varied with off-target regions but defaulted disproportionately to the reference allele. This default to the reference was additionally shown when counting both alleles at a locus where the reference allele was markedly outside of the distribution of alleles (teal is the reference allele; orange is the non-reference).

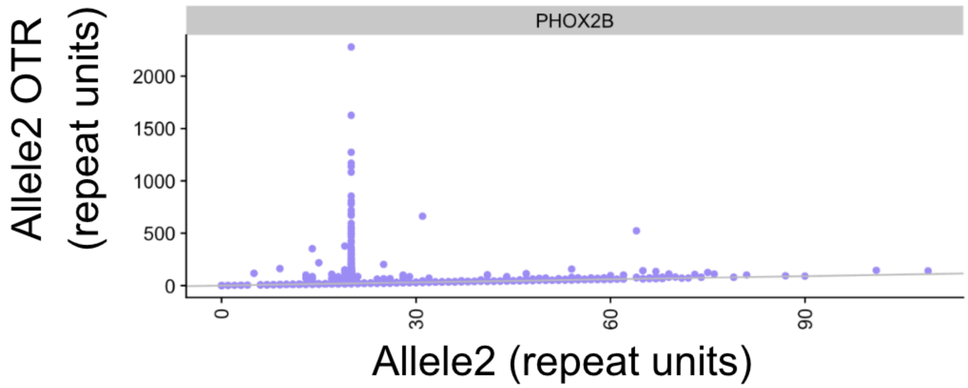


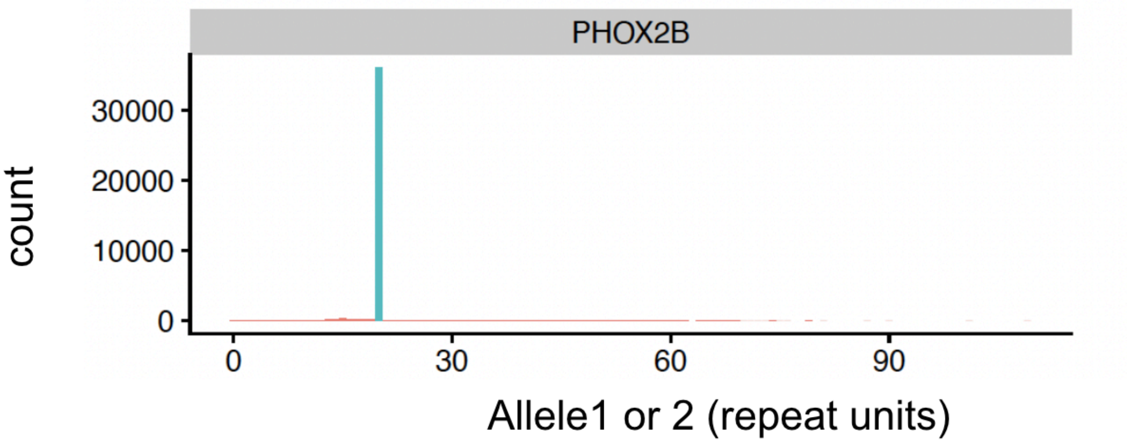


Additionally, when calculating the PGs and PG percentages for the CNG locus, the pathogenic percentage was substantially higher than expected for several congenital conditions (see Figure 3). All loci with CNG motifs had PGs except for females in the X-linked recessive conditions at *ARX_2* and *SOX3*. Of these 18 CNG regions, *FOXL2,* the three *HOXA13* loci, *HOXD13*, *RUNX2*, *TBX1*, *ZIC2*, and *ZIC3* are all congenital conditions presenting at birth, and all had PGs in this data set. In fact, the three *HOXA13* loci ranged from 7 to almost 17% for PG percent. Despite these being incredibly rare conditions that should be present in early childhood, there were also relatively high rates of ARX PGs. There are no PGs for the non-CNG congenital disease loci of *XYLT1*, *EIF4A3*, and *PRDM12* (*CBL* was not in the gnomAD analysis).

| | gene | Pathogenic_Count | Motif | Inheritance | Total_Loci | Carrier_Count | pathogenic_percent |
|---|---|---|---|---|---|---|---|
| | All | All | CN | All | All | All | All |
| 19 | HOXA13_2 | 3099 | CNG | AD | 18508 | NA | 16.74411065 |
| 18 | HOXA13_1 | 2235 | CNG | AD | 18508 | NA | 12.07585909 |
| 20 | HOXA13_3 | 1334 | CNG | AD | 18508 | NA | 7.20769397 |
| 36 | RUNX2 | 214 | CNG | AD | 18505 | NA | 1.15644420 |
| 31 | PHOX2B | 205 | CNG | AD | 18508 | NA | 1.10762913 |
| 62 | XY_ARX_1 | 69 | CNG | XR | 10521 | NA | 0.65583119 |
| 63 | XY_ARX_2 | 68 | CNG | XR | 10519 | NA | 0.64644928 |
| 66 | XY_ZIC3 | 68 | CNG | XR | 10569 | NA | 0.64339105 |
| 44 | ZIC2 | 84 | CNG | AD | 18508 | NA | 0.45385779 |
| 16 | FOXL2 | 61 | CNG | AD | 18505 | NA | 0.32964064 |
| 40 | TBX1 | 54 | CNG | AD | 18507 | NA | 0.29178149 |
| 65 | XY_SOX3 | 24 | CNG | XR | 10529 | NA | 0.22794187 |
| 59 | XX_ZIC3 | 11 | CNG | XR | 7904 | 121 | 0.13917004 |
| 30 | PABPN1 | 9 | CNG | AD | 18508 | NA | 0.04862762 |
| 55 | XX_ARX_1 | 1 | CNG | XR | 7904 | 136 | 0.01265182 |
| 21 | HOXD13 | 2 | CNG | AD | 18508 | NA | 0.01080614 |
| 56 | XX_ARX_2 | 0 | CNG | XR | 7904 | 204 | 0.00000000 |
| 58 | XX_SOX3 | 0 | CNG | XR | 7904 | 123 | 0.00000000 |

*PG results subset to CNG motif.*

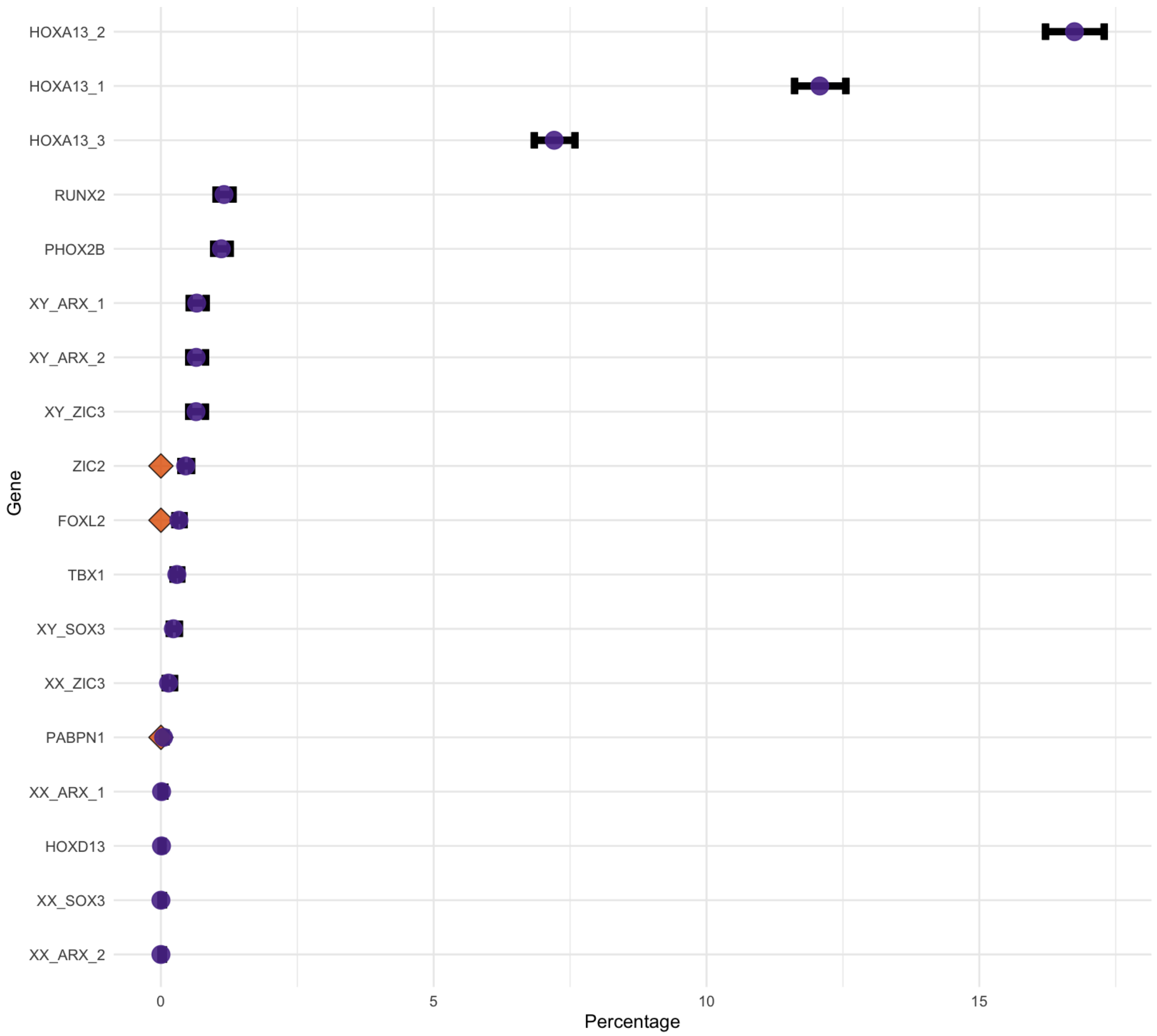


*CNG-specific percentages plotted.*

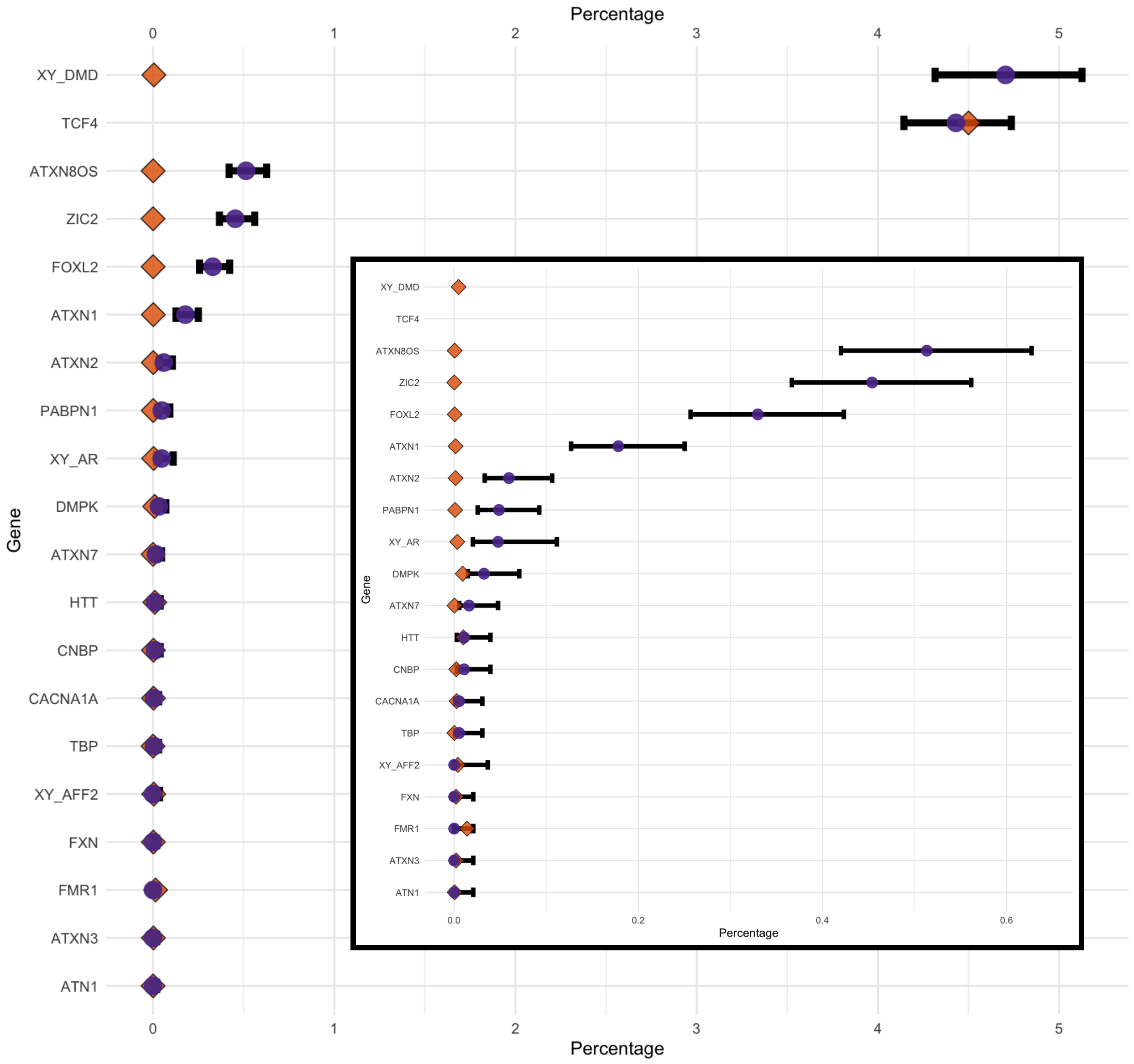


*CNGs are plotted alongside other motifs where disease prevalence is estimated.*

The non-specificity of CNG in the sequence may lead to inflated allele estimates outside of the reference genotyping errors. Additionally, five of these 18 loci are close together and coincide with multiple repetitive regions. This may lead to repeat conflation across reads and explain the exceptionally large PG percentage of the *HOXA13* loci.

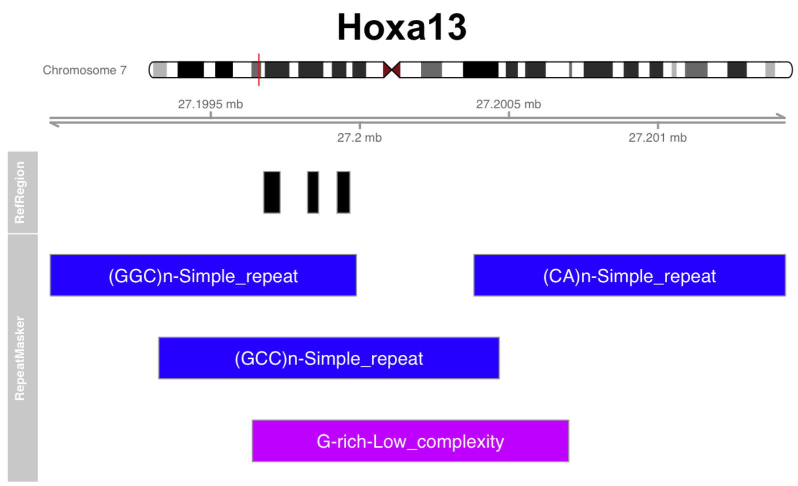


*STRchive* HOXA13 *loci plotted alongside RepeatMasker tracts in UCSC Genome Browser.*

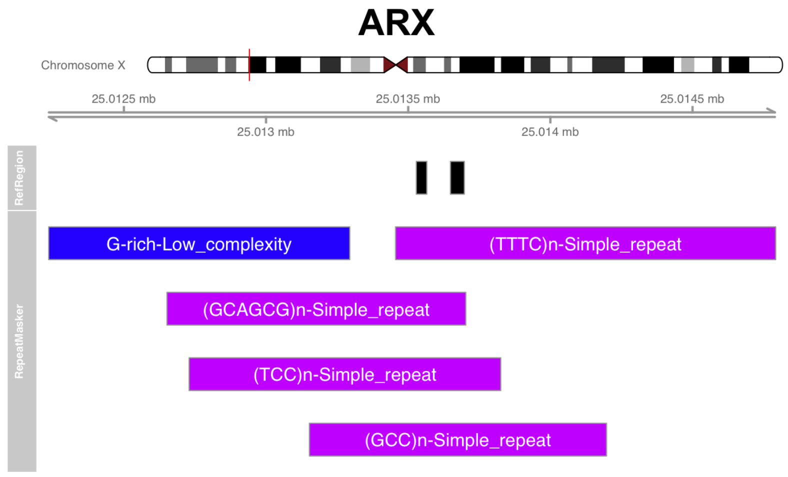


*STRchive* ARX *loci plotted alongside RepeatMasker tracts in UCSC Genome Browser.*

### Comparison with Tucci et al.

All code and logic for comparison are available within Github scripts folder in ComparisonwithTuccietal.R.

### Long-read data

Long-read data from the Human Pangenome Reference Consortium was provided in VCF format by co-author Egor Dolzhenko, genotyped by TRGT based on the TRGT variant catalog available on STRchive. Python file trgt_vcf_to_tsv.py was used to create two TSV files from the TRGT VCFs, one with allele lengths (HPRC.allele_lengths.tsv) and one with motif counts( HPRC.motif_counts.tsv).

*Motif analysis*

Compound loci were evaluated by the bash code:

```
awk '$6 ~ /n\(/ {print $5}' data/HPRC.allele_lengths.tsv | sort -u
```

And saved as “locus_id_with_compound_structure.txt”.

tr-solve_HPRC_100.py was utilized to parse the locus motifs from each respective VCF file, and tr-solve.sh was used to apply this Python script to all VCFs. The combined output( combined_tr-solve_output.tsv) was subset to calls with multiple motifs with the bash script:

```
awk '$3 ~ /,/' data/combined_tr-solve_output.tsv > comma_combined_output.tsv
```

The unique motifs were extracted by comma_combined_output.tsv with the Python script:

```
#Step 1: Import file and process to unique columns
import re
f = pd.read_csv('~/Git/STRchive_manuscript/data/comma_combined_output.tsv', sep='\t',
header = None)
unique_ids = f.iloc[:, 5].unique()


# Step 2: Iterate over unique IDs and extract motifs
for unique_id in unique_ids:
    filtered_df = f[f.iloc[:, 5] == unique_id]
    motifs_strings = filtered_df[2]

    unique_motifs = set()
    for motif_str in motifs_strings:
        motifs = re.findall(r"'([^']*)'", motif_str)
        unique_motifs.update(motifs)

    print(f"Unique motifs for ID {unique_id}:")
    print(unique_motifs)
    print("Total unique motifs:", len(unique_motifs))
```

This output was then compared against compound loci IDs to evaluate loci with unique motifs, which were then pasted as a tibble into Figure4.R to visualize the motif heterogeneity from the long-read data.

*DMD PG analysis*
DMD PG analysis was performed using the script long_read_PG.Rmd, which is identical to the primary PG code but excludes motif comparison, given the complexity of the motif output and its irrelevance to evaluating *DMD*. The PG percentage and specific counts were then taken from the subsequent combined_df.

The *DMD* allele sequences were then reviewed manually with the following code:

```
awk '$6 ~ /ID=DMD_DMD/' data/combined_tr-solve_output.tsv > DMD.tsv
```

When allele sequences for *DMD* were manually reviewed, all but one female were validated: one female had interruptions that reduced the number of complete repeats to 58 repeats, a level of sequence resolution unlikely to be caught by short-read sequencing (Supplementary Methods). Although *DMD* PGs remain far higher than expected overall, this borderline PG illustrates how pathogenic thresholds as static parameters may be challenged by different sequencing and genotyping methods that may inflate, deflate, or erase allelic attributes.

MC_vcf represented motif counts, two alleles for females and one for males) to confirm motif length (ALT sequence) with reported repeat counts from the genotype.

```
carriers
113 -> 112
HG02280
ALT=TTCTTCTTCTTCTTCTTCTTCTTCTTCTTCTTCTTCTTCTTCTTCTTCTTCTTCTTCTTCTTCTTCTTCTTCTTCTTCTTCTTCT
TCTTCTTCTTCTTCTTCTTCTTTTCTTCTTCTTCTTCTTCTTCTTCTTCTTCTTCTTCTTCTTCTTCTTCTTCTTCTTCTTCTTCTTCT
TCTTCTTCTTCTTCTTCTTCTTCTTCTTCTTCTTCTTCTTCTTCTTCTTCTTCTTCTTCTTCTTCTTCTTCTTCTTCTTCTTCTTCTTC
TTCTTCTTCTTCTTCTTCTTCTTCTTCTTCTTCTTCTTCTTCTTCTTCTTCTTCTTCTTCTTCTTCTTCTTCTTCTTCTTCTTCTTC
MOTIFS={'TTC'}  MOTIF=TTC   STRUC=(TTC)n    ID=DMD_DMD  MC_VCF=['22,113']
103 -> 100
HG02630
ALT=TTCTTCTTCTTCTTCTTCTTCTTCTTCTTCTTCTTCTTCTTCTTCTTCTTCTTCTTCTTCTTCTTCTTCTTCTTCTTTTCCT
CTTCTTCTTCTTCTTCTTCTTCTTCTTCTTCTTCTTCTTCTTCTTCTTCTTCTTCTTCTTCTTCTTCTTCTTCTTCTTCTTCTTCTTCT
TCTTCTTCTTCTTCTTCTTCTTCTTCTTCTTCTTCTTCTTCTTCTTCTTCTTCTTCTCTTCTTCTTCTTCTTCTTCTTCTTCTTCTTCT
TCTTCTTCTTCTTCTTCTTCTTCTTCTTCTCTTCTTCTTCTTCTTCTTCTTCTTC   MOTIFS={'TTC'}  MOTIF=TTC
STRUC=(TTC)n    ID=DMD_DMD  MC_VCF=['24,103']
72 -> 69
HG02976
ALT=TTCTTCTTCTTCTTCTTCTTCTTCTTCTTCCTTCTTTCTTCTTCTTCTTCTTCTTCTTCTTCTTCTTCTTCTTCTTCTTCTT
CTTCTTCTTCTTCTTCTTCTTCTTCTTCTTCTTCTTCTTCTTCTTCTTCTTCTTCTTCTTCTTCTTCTTCTTCTTCTTCTTCTTCTTCT
TCTTCTTCTTCTTCTTCTTCTTCTTCTTCTTCTTCTTCTTCTTT    MOTIFS={'TTC'}  MOTIF=TTC
STRUC=(TTC)n    ID=DMD_DMD  MC_VCF=['22,72']
85 -> 81
HG03041
ALT=TTTTTCTTCTTCTTCTTTCTTCTTCCTTCTTCTTCTTCTTCTTCTTCTTCTTCTTCTTCTTCTTCTTCTTCTTCTTCTTCTT
CTTCTTCTTCTTCTTCTTCTTCTTCTTCTTCTTCTTCTTCTTCTTCTTCTTCTTCTTCTTCTTCTTCTTCTTCTTCTTCTTCTTCTTCT
TCTTCTTCTTCTTCTTCTTCTTCTTCTTCTTCTTCTTCTTCTTCTTCTTCTTCTCTTCTTCTTCTTCTTCTTCTTCTTCTTCTTCTT
MOTIFS={'TTC'}  MOTIF=TTC   STRUC=(TTC)n    ID=DMD_DMD  MC_VCF=['23,85']
97 -> 76
HG03516
ALT=TTCTTCTTCTTCTTCTTCTTCTTCTTCTTCTTCTTCTTCTTCTTCTTCTTCTTCTTCTTCTTCTTCTTCTTCCTCTTCTTCT
```

```
TCTTCTTCCTCTTCCTCTTCCTCTTCCTCTTCCTCTTCCTCTTCCTCTTCCTCTTCCTCTTCCTCTTCCTCTTCCTCTTCCTCTTC
CTCTTCTTCTTCCTCTTCTTCTTCTTCTTCCTCTTCTTCTTCCTCTTCCTCTTCTTCTTCCTCTTCTTCTTCTTCTTCTTCTTCTT
CTTCTTCTTCTTCTTCTTCTTCTTCTTCTTCTTC    MOTIFS={'TTC'}  MOTIF=TTC   STRUC=(TTC)n
ID=DMD_DMD  MC_VCF=['12,97']
83 -> 82
HG03540
ALT=TTCTTCTTCTTCTTCTTCTTCTTCTTCTTCTTCTTCTTTCTTCTTCTTCTTCTTCTTCTTCTTCTTCTTCTTCTTCTTCTTC
TTCTTCTTCTTCTTCTTCTTCTTCTTCTTCTTCTTCTTCTTCTTCTTCTTCTTCTTCTTCTTCTTCTTCTTCTTCTTCTTCTTCTT
CTTCTTCTTCTTCTTCTTCTTCTTCTTCTTCTTCTTCTTCTTCTTCTTCTTCTTCTTCTTCTTCTTCTTCTTCTTCTTCTTC
MOTIFS={'TTC'}  MOTIF=TTC   STRUC=(TTC)n    ID=DMD_DMD  MC_VCF=['13,83']


males
102 -> 99
HG03579
ALT=TTTTTCTTCTTCTTCTTTCTTCTTCCTTCTTCCTTCTTCTTCTTCTTCTTCTTCTTCTTCTTCTTCTTCTTCTTCTTCTTCT
TCTTCTTCTTCTTCTTCTTCTTCTTCTTCTTCTTCTTCTTCTTCTTCTTCTTCTTCTTCTTCTTCTTCTTCTTCTTCTTCTTCTTC
TTCTTCTTCTTCTTCTTCTTCTTCTTCTTCTTCTTCTTCTTCTTCTTCTTCTTCTTCTTCTTCTTCTTCTTCTTCTTCTTCTTCTT
CTTCTTCTTCTTCTTCTTCTTCTTCTTCTTCTTCTTCTTCTTCTTCTTC MOTIFS={'TTC'}  MOTIF=TTC
STRUC=(TTC)n    ID=DMD_DMD  MC_VCF=['102']
91 -> 87
NA19043
ALT=TTTTTCTTCTTCTTCTTTCTTCTTCCTTCTTCTTCCTTCTTCCTTCTTCTTCTTCTTCTTCTTCTTCTTCTTCTTCTTCTTC
TTCTTCTTCTTCTTCTTCTTCTTCTTCTTCTTCTTCTTCTTCTTCTTCTTCTTCTTCTTCTTCTTCTTCTTCTTCTTCTTCTTCTT
CTTCTTCTTCTTCTTCTTCTTCTTCTTCTTCTTCTTCTTCTTCTTCTTCTTCTTCTTCTTCTTCTTCTTCTTCTTCTTCTTCTTCT
TCTTCTTCTTCTTC    MOTIFS={'TTC'}  MOTIF=TTC   STRUC=(TTC)n    ID=DMD_DMD
MC_VCF=['91']

females
84 -> 84
HG02559
ALT=TTCTTCTTCTTCTTCTTCTTCTTCTTCTTCTTCTTCTTCTTCTTCTTCTTCTTCTTCTTCTTCTTCTTCTTCTTCTTCTTCT
TCTTCTTCTTCTTCTTCTTCTTCTTCTTCTTCTTCTTCTTCTTCTTCTTCTTCTTCTTCTTCTTCTTCTTCTTCTTCTTCTTCTTC
TTCTTCTTCTTCTTCTTCTTCTTCTTCTTCTTCTTCTTCTTCTTCTTCTTCTTCTTCTTCTTCTTCTTCTTCTTCTTCTTCTTC
MOTIFS={'TTC'}  MOTIF=TTC   STRUC=(TTC)n    ID=DMD_DMD  MC_VCF=['84,117']
117 -> 113
HG02559
ALT=TTCTTCTTCTTCTTCTTCTTCTTCTTCCTTCTTCTTCTTCTTCTTCTTCTTCTTCTTCTTCTTCTTCTTCTTCTTCTTCTTC
TTCTTCTTCTTCTTCTTCTTCTTCTTCTTCTTCTTCTTCTTCTTCTTCTTCTTCTTCTTCTTCTTCTTCTTCTTCTTCTTCTTCTT
CTTCTTCTTCTTCTTCTTCTTCTTCTTCTTCTTCTTCTTCTTCTTCTTCTTTTTCTTCTTCTTCTTCTTCTTCTTCTTCTTCTTCT
TCTTCTTCTTCTTCTTCTTCTTTCTTCTTCTTCTTCTTCTTCTTCTTCTTCTTCTTCCTTCTTCTTCTTCTTCTTCTTCTTCT
TCTCTTC MOTIFS={'TTC'}  MOTIF=TTC   STRUC=(TTC)n    ID=DMD_DMD  MC_VCF=['84,117']
```

```
147 -> 143
HG02622
ALT=TTCTTCTTCTTCTTCTTCTTCTTCCTTCTTCTTCTCTTTCTTCTTCTTCTTCTTCTTCTTCTTCTTCTTCTTCTTCTTCTTC
TTCTTCTTCTTCTTCTTCTTCTTCTTCTTCTTCTTCTTCTTCTTCTTCTTCTTCTTCTTCTTCTTCTTCTTCTTCTTCTTCTTCTT
CTTCTTCTTCTTCTTCTTCTTCTTCTTCTCTTCTTCTTCTTCTTCTTCTTCTTCTTCTCTTCTTCTTCTTCTTCTTCTTCTTCTTC
TTCTTCTTCTTCTTTCTTCTTCTTCTTCTTCTTCTTCTTCTTCTTCTTCTTCTTCTTCTTCTTCTTCTTCTTCTTCTTCTTCTTCT
TCTTCTTCTTCTTCTTCTTCTTCTTCTTCTTCTTCTTCTTCTTCTTCTTCTTCTTCTTCTTCTTCTTCTTCTTCTTCTTCTTCTTC
TTCTTCTTCTTC   MOTIFS={'TTC'}  MOTIF=TTC    STRUC=(TTC)n     ID=DMD_DMD
MC_VCF=['59,147']
61 -> 58
HG02622
ALT=TTCTTCTTCTTCTTCTTCTTCTTCTTCTTCTTCTTCTTCTTCTTCTTCTTCTTCTTCTTCTTCTTCTTCTTCTTCTTCTTCTTTT
CTTCTTCTTCTTCTTCTTCTTCTTCTTCTTCTTCTTCTTCTTCTTCTTCTTCTTCTTCTTCTTCTTCTTCTTCTTCTTCTTCTTCT
TCTTCTTC     MOTIFS={'TTC'}  MOTIF=TTC    STRUC=(TTC)n     ID=DMD_DMD  MC_VCF=['59,147']
```

### Catalog Differences with gnomAD

The following output was generated by CatalogDifferences.ipynb to compare the gnomAD variant catalog to the STRchive catalog, with differences of more than 2bp on either end bolded and annotations added. Twenty-seven loci were identical between catalogs (italicized).

Gene: ABCD3, Start Difference: 1, Stop Difference: 2
**Gene: AFF2, Start Difference: -26, Stop Difference: 62 *TRF in UCSC Browser differs/extends past gnomAD definition**
Gene: AFF3, Start Difference: 1, Stop Difference: 2
**Gene: AR, Start Difference: 1, Stop Difference: 34 *TRF in UCSC Browser extends past gnomAD definition**
*Gene: ARX, Start Difference: 0, Stop Difference: 0*
Gene: ARX_2, Start Difference: 1, Stop Difference: 0
Gene: ATN1, Start Difference: 1, Stop Difference: 2
Gene: ATXN1, Start Difference: 1, Stop Difference: 1
Gene: ATXN10, Start Difference: 1, Stop Difference: 0
Gene: ATXN2, Start Difference: 1, Stop Difference: 1
**Gene: ATXN3, Start Difference: 2, Stop Difference: 13 *TRF in UCSC Browser extends past gnomAD definition**
Gene: ATXN7, Start Difference: 1, Stop Difference: 2
**Gene: BEAN1, Start Difference: -1, Stop Difference: 13 *TRF in UCSC Browser extends past gnomAD definition**
**Gene: C9orf72, Start Difference: -44, Stop Difference: 0 *TRF in UCSC Browser extends past gnomAD definition**
Gene: CACNA1A, Start Difference: 1, Stop Difference: 1
*Gene: CBL, Start Difference: 0, Stop Difference: 0*
Gene: CNBP, Start Difference: 1, Stop Difference: 3
Gene: COMP, Start Difference: 0, Stop Difference: 0
Gene: CSTB, Start Difference: 0, Stop Difference: 0
**Gene: DAB1, Start Difference: 1, Stop Difference: 7 *TRF in UCSC Browser extends past gnomAD definition**

Gene: DIP2B, Start Difference: 1, Stop Difference: 0
*Gene: DMD, Start Difference: 0, Stop Difference: 0*
Gene: DMPK, Start Difference: 0, Stop Difference: 2
Gene: EIF4A3, Start Difference: 0, Stop Difference: 0
**Gene: FGF14, Start Difference: 3, Stop Difference: 2 *TRF in UCSC Browser extends past gnomAD definition**
**Gene: FMR1, Start Difference: -13, Stop Difference: 1 *TRF in UCSC Browser extends past gnomAD definition, taken from literature consensus**
*Gene: FOXL2, Start Difference: 0, Stop Difference: 0*
*Gene: FXN, Start Difference: 0, Stop Difference: 0*
*Gene: GIPC1, Start Difference: 0, Stop Difference: 0*
Gene: GLS, Start Difference: 1, Stop Difference: 0
*Gene: HOXA13, Start Difference: 0, Stop Difference: 0*
*Gene: HOXA13_2, Start Difference: 0, Stop Difference: 0*
*Gene: HOXA13_3, Start Difference: 0, Stop Difference: 0*
*Gene: HOXD13, Start Difference: 0, Stop Difference: 0*
**Gene: HTT, Start Difference: 1, Stop Difference: 7 *TRF in UCSC Browser extends past gnomAD definition**
**Gene: JPH3, Start Difference: -4, Stop Difference: 0 *TRF in UCSC Browser extends past gnomAD definition**
Gene: LRP12, Start Difference: 2, Stop Difference: 2
**Gene: MARCHF6, Start Difference: -7, Stop Difference: 0 *TRF in UCSC Browser extends past gnomAD definition**
*Gene: NIPA1, Start Difference: 0, Stop Difference: 0*
*Gene: NOP56, Start Difference: 0, Stop Difference: 0*
Gene: NOTCH2NLC, Start Difference: 1, Stop Difference: 1
**Gene: PABPN1, Start Difference: 0, Stop Difference: 12 *TRF in UCSC Browser extends past gnomAD definition**
*Gene: PHOX2B, Start Difference: 0, Stop Difference: 0*
Gene: PPP2R2B, Start Difference: 1, Stop Difference: 2
*Gene: PRDM12, Start Difference: 0, Stop Difference: 0*
*Gene: PRNP, Start Difference: 0, Stop Difference: 0*
Gene: RAPGEF2, Start Difference: 1, Stop Difference: 2
**Gene: RFC1, Start Difference: 1, Stop Difference: 4 *TRF in UCSC Browser extends past gnomAD definition**
**Gene: RILPL1, Start Difference: 0, Stop Difference: 5 *TRF in UCSC Browser extends past gnomAD definition**
*Gene: RUNX2, Start Difference: 0, Stop Difference: 0*
Gene: SAMD12, Start Difference: 1, Stop Difference: 0
*Gene: SOX3, Start Difference: 0, Stop Difference: 0*
Gene: STARD7, Start Difference: 1, Stop Difference: 0
Gene: TBP, Start Difference: 1, Stop Difference: 0
*Gene: TBX1, Start Difference: 0, Stop Difference: 0*
*Gene: TCF4, Start Difference: 0, Stop Difference: 0*
*Gene: THAP11, Start Difference: 0, Stop Difference: 0*
**Gene: TNRC6A, Start Difference: 0, Stop Difference: 3 *TRF in UCSC Browser extends past gnomAD definition**
*Gene: VWA1, Start Difference: 0, Stop Difference: 0*
*Gene: XYLT1, Start Difference: 0, Stop Difference: 0*
Gene: YEATS2, Start Difference: 1, Stop Difference: 0
Gene: ZFHX3, Start Difference: 1, Stop Difference: 1
*Gene: ZIC2, Start Difference: 0, Stop Difference: 0*
*Gene: ZIC3, Start Difference: 0, Stop Difference: 0*
Gene: ZNF713, Start Difference: 1, Stop Difference: 0
