## Supplementary Figures & Tables for "STRchive: a dynamic resource detailing population-level and locus-specific insights at tandem repeat disease loci"

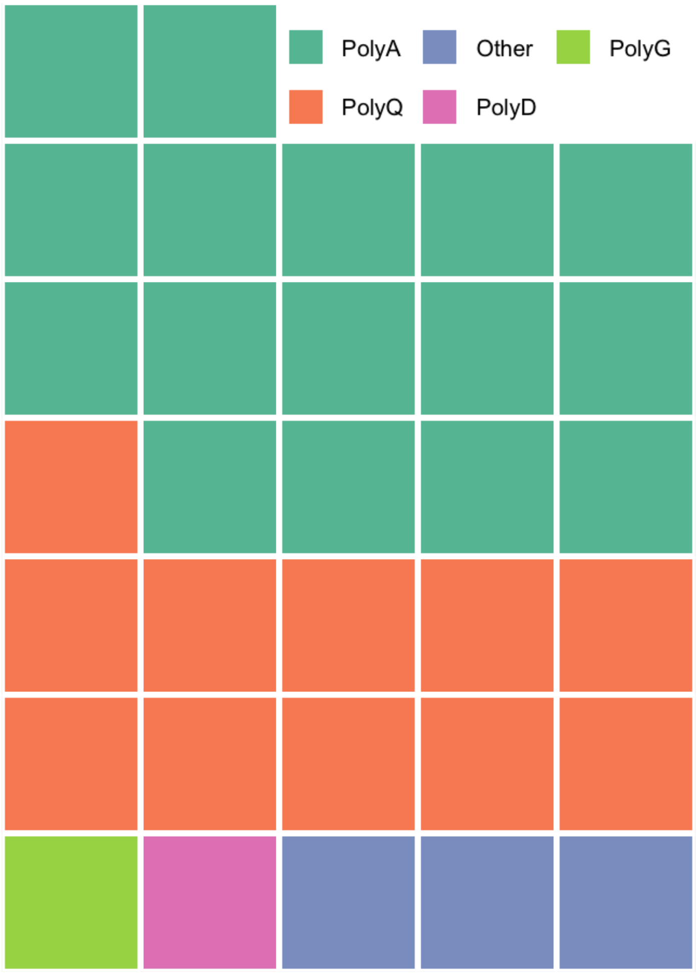

**Supplementary Figure 1: The majority of coding TRs result in polyalanine and polyglutamine tracts.** *A waffle plot of the coding types' specific coding consequence. Script for generation (with further details of the other categories) is available in Figure 1 script.*

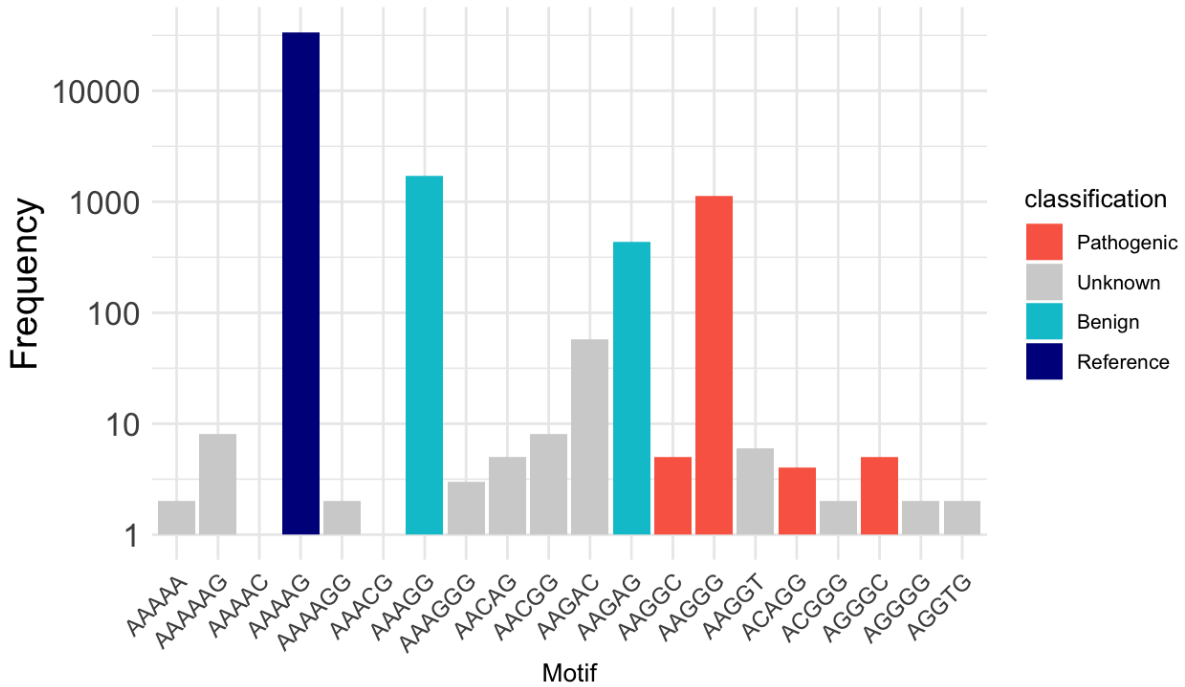

**Supplementary Figure 2: RFC1 has the highest motif diversity of the gnomAD dataset, with motifs of all classifications.** *RFC1 motif counts from the gnomAD data for all motifs, showing a relatively high proportion of pathogenic motifs as well as a collectively high proportion of unknown motifs. Script is available in script for Figure 5 generation.*

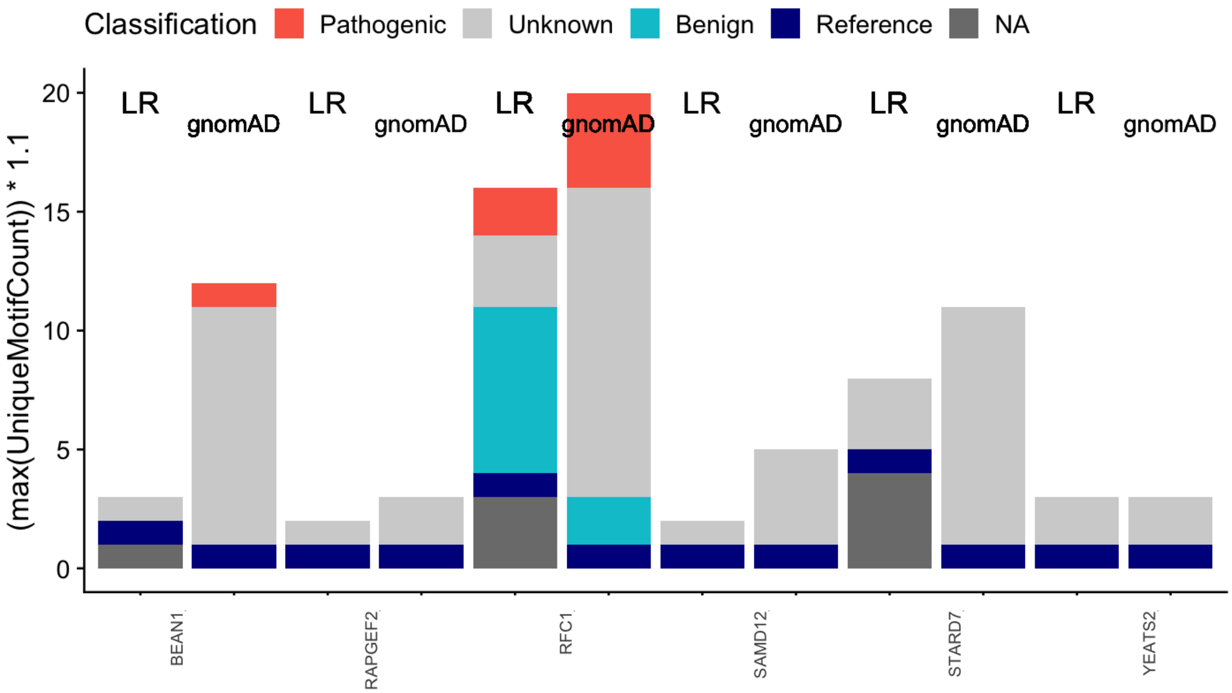

**Supplementary Figure 3: Six TR disease loci show motif heterogeneity across gnomAD and HPRC cohorts.** *Unique motifs taken from the HPRC long-read data, classified by STRchive, compared to the gnomAD data at overlapping loci. NA indicates "Not Available" in STRchive; other "Unknown" motifs have been documented in the literature before despite not being associated with a phenotype. Script is available in script for Figure 5 generation.*

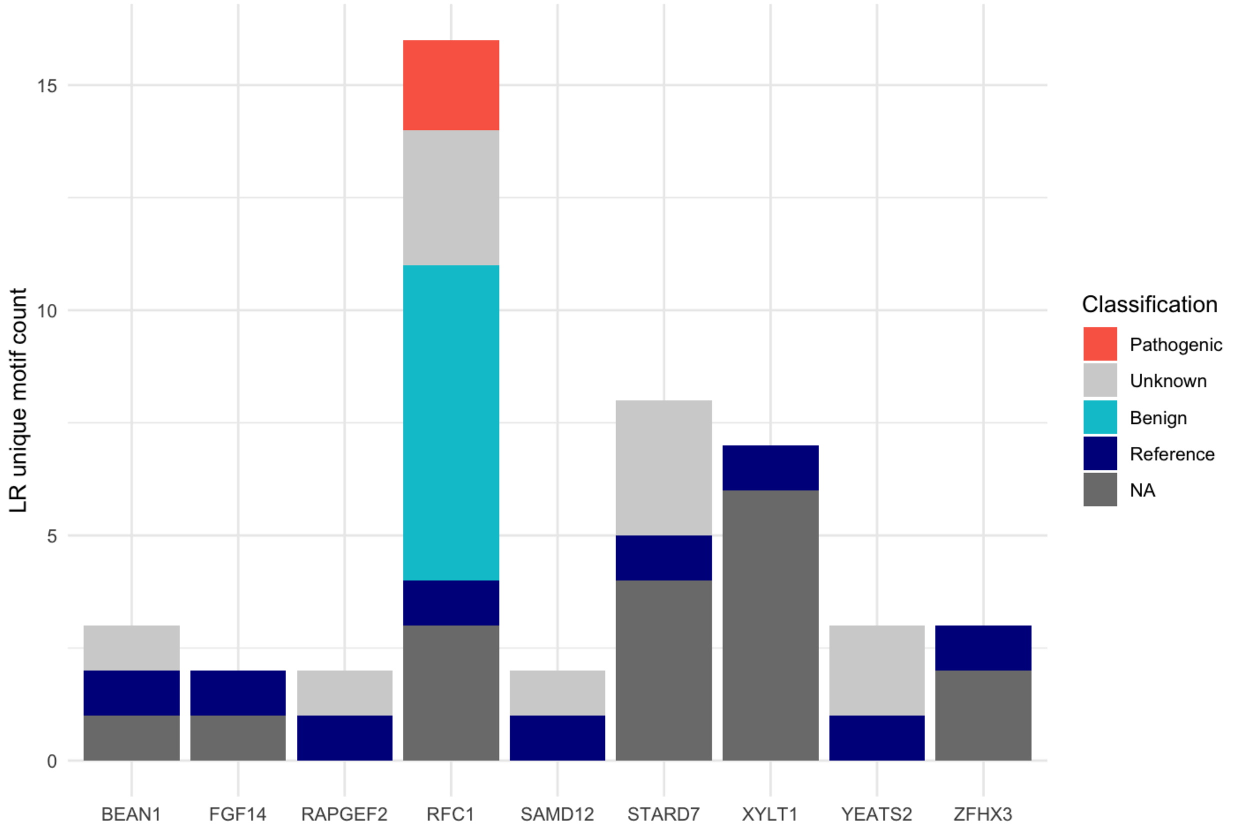

**Supplementary Figure 4: Nine loci show motif heterogeneity within HPRC cohort, with six having motifs previously not associated within individual genotypes.** *Unique motifs taken from the HPRC long-read data, classified by STRchive, for all loci with more than one unique motif. NA indicates "Not Available" in STRchive; other "Unknown" motifs have been documented in the literature before despite not being associated with a phenotype. Script is available in script for Figure 5 generation.*

**Supplementary Table 1: PGs are calculated at gnomAD TR disease loci and compared to disease prevalence, where known.** *All gnomAD loci where PGs were calculated, with PG percentage, carrier percentage, prevalence, and 95% binomial confidence interval shown. All numbers are rounded to four decimal places of percentage value. Data generated by CalculatingPGsandConfidenceIntervals.R.*

| Gene (AD) | gnomAD PG % (95% CI) | Disease Prevalence % |
|---|---|---|
| *TCF4* | 4.4305 (4.1434–4.7365) | 4.5000 |
| *ATXN8OS* | 0.5133 (0.4201–0.627) | 0.0005 |
| *PRNP* | 0.3188 (0.2472–0.4109) | *NA* |
| *NIPA1* | 0.281 (0.2144–0.3683) | *NA* |
| *ATXN1* | 0.1783 (0.127–0.2503) | 0.0015 |
| *ATXN2* | 0.0594 (0.0332–0.1064) | 0.0015 |
| *PABPN1* | 0.0486 (0.0256–0.0924) | 0.0010 |
| *DMPK* | 0.0324 (0.0149–0.0707) | 0.0093 |
| *GIPC1* | 0.027 (0.0115–0.0632) | *NA* |
| *ATXN7* | 0.0162 (0.0055–0.0477) | 0.0003 |
| *HTT* | 0.0108 (0.003–0.0394) | 0.0100 |
| *CNBP* | 0.0108 (0.003–0.0394) | 0.0023 |
| *HOXD13* | 0.0108 (0.003–0.0394) | *NA* |
| *JPH3* | 0.0108 (0.003–0.0394) | *NA* |
| *CACNA1A* | 0.0054 (0.001–0.0306) | 0.0027 |
| *COMP* | 0.0054 (0.001–0.0306) | *NA* |
| *TBP* | 0.0054 (0.001–0.0306) | 0.0002 |
| *ATN1* | 0 (0–0.0208) | 0.0005 |
| *ATXN10* | 0 (0–0.0208) | *NA* |
| *ATXN3* | 0 (0–0.0208) | 0.0021 |
| *BEAN1* | 0 (0–0.0208) | *NA* |
| *C9ORF72* | 0 (0–0.0208) | *NA* |
| *DAB1* | 0 (0–0.0208) | *NA* |
| *DIP2B* | 0 (0–0.0208) | *NA* |
| *LRP12* | 0 (0–0.0208) | *NA* |
| *MARCHF6* | 0 (0–0.0208) | *NA* |
| *NOP56* | 0 (0–0.0208) | *NA* |

| | | |
|---|---|---|
| *NOTCH2NLC* | 0 (0–0.0208) | *NA* |
| *NUTM2B-AS1* | 0 (0–0.0208) | *NA* |
| *PPP2R2B* | 0 (0–0.0208) | *NA* |
| *RAPGEF2* | 0 (0–0.0208) | *NA* |
| *RILPL1* | 0 (0–0.0208) | *NA* |
| *SAMD12* | 0 (0–0.0208) | *NA* |
| *STARD7* | 0 (0–0.0208) | *NA* |
| *TNRC6A* | 0 (0–0.0208) | *NA* |
| *YEATS2* | 0 (0–0.0208) | *NA* |

| Gene (AR) | gnomAD Carrier % (95% CI) | gnomAD PG % (95% CI) | Disease Prevalence % |
|---|---|---|---|
| *FXN* | 0.2648 (0.2003–0.3498) | 0 (0–0.0208) | 0.0020 |
| *VWA1* | 0.1189 (0.0785–0.1799) | 0 (0–0.0208) | *NA* |
| *PRDM12* | 0.0432 (0.0219–0.0853) | 0 (0–0.0208) | *NA* |
| *XYLT1* | 0.0108 (0.003–0.0394) | 0 (0–0.0208) | *NA* |
| *CSTB* | 0.0054 (0.001–0.0306) | 0 (0–0.0208) | *NA* |
| *GLS* | 0.0054 (0.001–0.0306) | 0 (0–0.0208) | *NA* |
| *EIF4A3* | 0 (0–0.0208) | 0 (0–0.0208) | *NA* |
| *RFC1* | 0 (0–0.0208) | 0 (0–0.0208) | *NA* |

| Gene (XR) | Sex | gnomAD Carrier % (95% CI) | gnomAD PG % (95% CI) | Disease Prevalence % |
|---|---|---|---|---|
| *DMD* | XY | | 4.7048 (4.3161–5.1266) | 0.0048 |
| *DMD* | XX | 8.1984 (7.6137–8.8237) | 0.0886 (0.0429–0.1827) | *NA* |
| *AR* | XY | | 0.0477 (0.0204–0.1115) | 0.0033 |
| *AR* | XX | 0.0633 (0.027–0.148) | 0 (0–0.0486) | *NA* |
| *AFF2* | XY | | 0 (0–0.0365) | 0.004 |
| *AFF2* | XX | 0 (0–0.0486) | 0 (0–0.0486) | *NA* |

| Gene (XD) | gnomAD PG % (95% CI) | Disease Prevalence % |
|---|---|---|
| *FMR1* | 0 (0–0.0208) | 0.014 |

**Supplementary Table 2: Within gnomAD, PGs are found within 17 autosomal dominant loci and three X-linked recessive loci.** *gnomAD loci where at least one PG was found, with PG percentage and 95% binomial confidence interval shown in contrast with prevalence. All numbers are rounded to four decimal places of percentage value. Literature prevalence is bolded where it is within the confidence interval of the gnomAD pathogenic genotype (PG) percentage. For all other loci (including all AR and XD loci), prevalence was either unavailable or in the range of $10^{-(4\text{-}6)}$. Data generated by CalculatingPGsandConfidenceIntervals.R.*

| Gene (AD) | gnomAD PG % (95% CI) | Disease Prevalence % |
|---|---|---|
| *TCF4* | 4.4305 (4.1434–4.7365) | **4.5** |
| *ATXN8OS* | 0.5133 (0.4201–0.6270) | 0.0005 |
| *ATXN1* | 0.1783 (0.1270–0.2503) | 0.0015 |
| *ATXN2* | 0.0594 (0.0332–0.1064) | 0.0015 |
| *PABPN1* | 0.0486 (0.0256–0.0924) | 0.001 |
| *DMPK* | 0.0324 (0.0149–0.0707) | 0.008 |
| *ATXN7* | 0.0162 (0.0055–0.0477) | 0.0003 |
| *HTT* | 0.0108 (0.0030–0.0394) | **0.01** |
| *JPH3* | 0.0108 (0.0030–0.0394) | 0.0023 |
| *CAC1NAA* | 0.0054 (0.0010–0.0306) | 0.0003 |
| *TBP* | 0.0054 (0.0010–0.0306) | 0.0002 |
| *PRNP* | 0.3188 (0.2472–0.4109) | *NA* |
| *NIPA1* | 0.2810 (0.2144–0.3683) | *NA* |
| *GIPC1* | 0.0270 (0.0115–0.0632) | *NA* |
| *CNBP* | 0.0108 (0.0030–0.0394) | *NA* |
| *HOXD13* | 0.0108 (0.0030–0.0394) | *NA* |
| *COMP* | 0.0054 (0.0010–0.0306) | *NA* |

| Gene (XR) | Sex | gnomAD PG % (95% CI) | Disease Prevalence % |
|---|---|---|---|
| *DMD* | XY | 4.7048 (4.3161–5.1266) | 0.0048 |
| *DMD* | XX | 0.0886 (0.0429–0.1827) | *NA* |
| *AR* | XY | 0.0477 (0.0204–0.1115) | 0.0033 |

**Supplementary Table 3: A comprehensive blueprint identifying current resources relevant to each point is available on STRchive.**

| OVERVIEW | TR SPECIFIC DETAILS | | | RESOURCES |
|---|---|---|---|---|
| **Evaluating allele(s)** | | | | |
| Allele Size | *Premutations* | *Contraction/expansion* | *Somatic mosaicism* | |
| Compare allele of interest to available thresholds for benign, intermediate, and pathogenic size. | Evaluate whether an allele may be classified within the intermediate range as a premutation--this may have implications for patient presentation (mild or atypical phenotype) or for family members. | While most TR diseases are caused by expansions, contractions are speculated to lead to disease in specific loci where the reference allele is highly constrained. Consider whether an allele may be a pathogenic contraction versus an expansion. | Allelic instability may be tissue-specific; evaluate the sampled tissue and whether an allele may be a pathogenic size in the relevant tissue if the allele approaches a pathogenic threshold. | STRchive |
| Sequence Composition | *Motif classification* | *Interruptions* | | |
| Contrast allele sequence with reference and known TR sequences. | Determine if genotyped motif is benign, pathogenic, or of unknown consequence. | Assess motif sequence purity, as interruptions may increase or decrease penetrance, disease severity, or age of onset. | | STRchive, Rajan-Babu et al., 2024. |
| Genotype quality | *Read visualization* | *Experimental workflow* | | |
| Check genotype quality and read support to filter unreliable calls. | Review read visualizations for alleles of similar size to assess expected read support and pattern of interruptions. | Appraise the molecular and sequencing technologies used to identify the allele and how this may impact call reliability. | | gnomAD, Tanudisastro et al., 2024, Chaisson et al., 2023 |
| Allele frequency | *Ancestry-specific* | *Polymorphic distribution* | | |
| Determine allele frequency within broader population; variant frequency is likely to parallel disease penetrance and prevalence. | Populations with different ancestries may have different allelic distributions and thresholds for pathogenicity; review the allele in the context of | Given the highly polymorphic aspect of TRs, there are far more alleles likely to be present in a population at most loci than variants such as SNVs. As this may deflate exact allelic frequency, consider whether the allele falls outside of the normal distribution of alleles in addition to its exact frequency. | | gnomAD, WebSTR, STRipy, TRGT database |

| | | | | |
|---|---|---|---|---|
| | the relevant population if possible. | | | |
| Inheritance pattern | *Mixed mutation types* | | | |
| Assess both alleles (if present) in case of recessive condition. | Consider non-TR, potentially compounding variants in second allele. | | | STRchive, ClinVar, ClinGen |
| **Evaluating phenotype** | | | | |
| Genotype-phenotype correlation | *Anticipation* | *Reduced penetrance* | *Atypical presentation* | |
| Compare clinical history to symptoms associated with gene (if any). | TR diseases may demonstrate anticipation, where disease severity increases and age of onset decreases by generation as alleles expand through transmission. Consider family history. | Penetrance can vary due to genetic modifiers and allelic attributes (motif, interruptions, etc.). Recall that a pathogenic genotype may not indicate current or future disease. | TR disease can present with immensely variable phenotypes, both in terms of severity and specific symptoms. Often, there is an inverse correlation between allele size and age of onset, which can lead to early and late-onset diseases outside of the conventional range. | OMIM, GeneReviews, Orphanet |
| Assess whether patient history matches known interval of disease onset. | | | | STRchive |
| **Evaluating the locus** | | | | |
| Known disease association | *Predicted pathogenicity* | | | |
| Evaluate whether the locus has established association with TR disease by comparing to current catalogs. | There are loci that, while not associated with documented disease, have been predicted to be pathogenic through machine learning-based predictions. Additionally, manual comparison to known disease loci can inform the prediction of pathogenicity at novel loci based on known mechanisms of disease (e.g., polyalanine/glutamine tracts.) | | | STRchive, RExPRT |
| Identify whether the gene has previous gene-disease associations | | | | ClinGen, OMIM, GenCC |

| | | | | |
|---|---|---|---|---|
| documented for non-TR variant types. | | | | |
| Genomic region | *Proximity to another TR locus* | | | |
| The genomic region in which a locus is present is highly informative: whether coding/non-coding, whether it overlaps genetic elements such as promoters/enhancers, and whether nearby variants have known disease relevance. | Several TR disease loci are found within the same gene. TR locus proximity may indicate potential pathogenicity, but also may lead to inflated allele estimation. Leverage nearby loci to inform variant interpretation. | | | STRchive, ClinVar, UCSC Genome Browser |
| *The complexity and heterogeneity of TR loci means clinical and biological information may not be available in all cases. We recommend reviewing pertinent literature (cataloged by STRchive) and using best judgment when prioritizing variants.* | | | | |

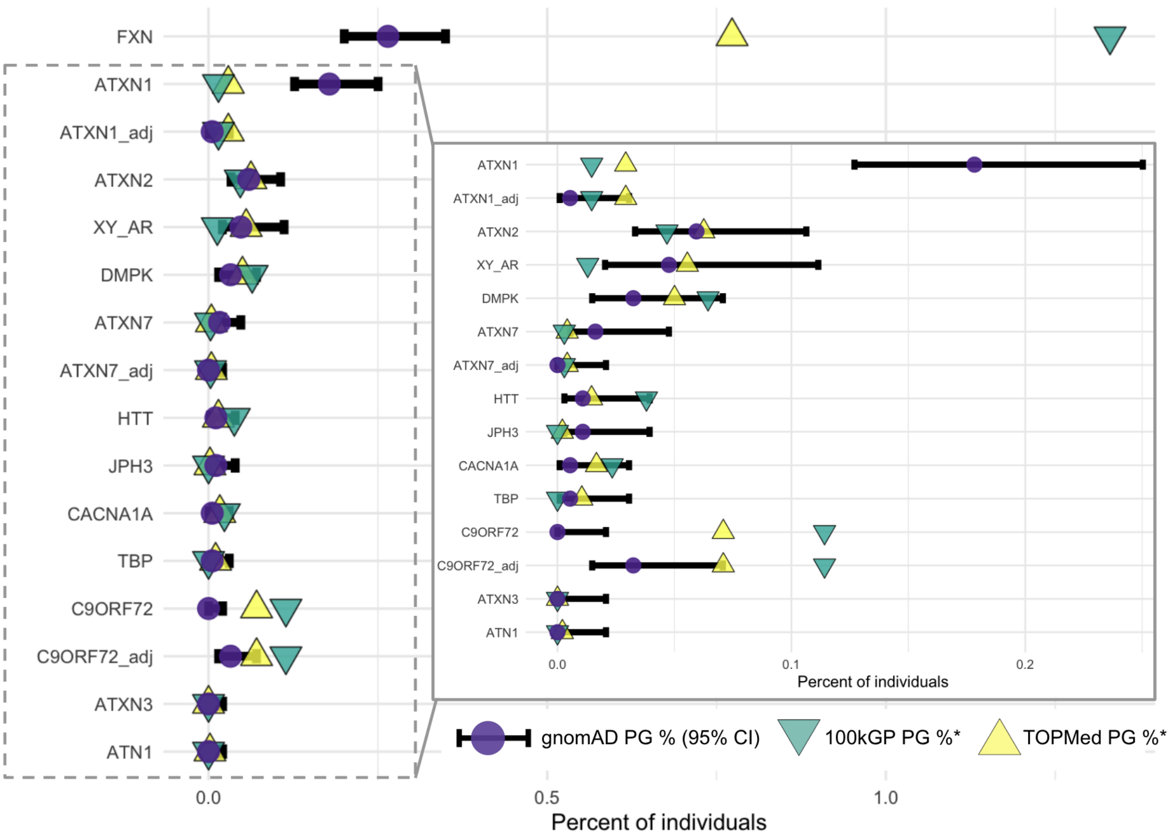

**Supplementary Figure 5: Comparisons between gnomAD, TOPMed, and 100kGP show general concordance in PG percentage.** An asterisk for PG % is used because *FXN* is an autosomal recessive locus, and so carrier percentage is used across cohorts rather than pathogenic genotypes. Matching the pathogenic thresholds in gnomAD to what was used by Tucci et al. for three non-equivalent loci (*ATXN1, ATXN7, C9ORF72*) is indicated by *_adj. This adjustment altered the PG confidence interval to span the TOPMed/100kGP PG percentages for the first two loci while adjusting the pathogenic threshold for *C9ORF72* brought the TOPMed within 0.00018798% of the upper limit of the 95% confidence interval. The TOPMed estimate for *JPH3* was within 0.000879498% of the lower limit of the 95% confidence interval. Curiously, the gnomAD *FXN* PG percentage is comparable to disease prevalence in the literature despite being 37 to 67-fold lower than TOPMed/100kGP estimates. The *FXN* analysis within TOPMed and 100kGP shows ancestry-based PG variation, which aligns with variation in prevalence estimates across different populations reported previously for *FXN*.[71] Consequently, the extreme variation in *FXN* PG percentages across studies may be due to cohort-specific ancestries.

| gene | percentage_100kGP | percentage_TOPMed | pathogenic_percent | Pathogenic_Count_lower_ci | Pathogenic_Count_upper_ci | Inheritance |
|---|---|---|---|---|---|---|
| ATN1 | 0.000000000 | 0.002083985 | 0.000000000 | 0.0000000000 | 0.02075360 | AD |
| ATXN1 | 0.014626298 | 0.029183690 | 0.178301275 | 0.1269935028 | 0.25028641 | AD |
| ATXN1_adj | 0.014626298 | 0.029183690 | 0.005403069 | 0.0009537807 | 0.03060147 | AD |
| ATXN2 | 0.046819219 | 0.062557344 | 0.059436970 | 0.0331929258 | 0.10640883 | AD |
| ATXN3 | 0.000000000 | 0.000000000 | 0.000000000 | 0.0000000000 | 0.02075136 | AD |
| ATXN7 | 0.002924917 | 0.004168056 | 0.016209207 | 0.0055127427 | 0.04765030 | AD |
| ATXN7_adj | 0.002924917 | 0.004168056 | 0.000000000 | 0.0000000000 | 0.02075136 | AD |
| C9ORF72 | 0.114198706 | 0.070904238 | 0.000000000 | 0.0000000000 | 0.02075136 | AD |
| C9ORF72_adj | 0.114198706 | 0.070904238 | 0.032418414 | 0.0148584698 | 0.07071626 | AD |
| CACNA1A | 0.023404131 | 0.016674309 | 0.005403653 | 0.0009538838 | 0.03060478 | AD |
| DMPK | 0.064387731 | 0.050039615 | 0.032418414 | 0.0148584698 | 0.07071626 | AD |
| FXN | 1.330798479 | 0.772817002 | 0.264750378 | 0.2003322211 | 0.34981002 | AR |
| HTT | 0.038037277 | 0.014589716 | 0.010806722 | 0.0029636429 | 0.03939780 | AD |
| JPH3 | 0.000000000 | 0.002083985 | 0.010806138 | 0.0029634828 | 0.03939567 | AD |
| TBP | 0.000000000 | 0.010420792 | 0.005403069 | 0.0009537807 | 0.03060147 | AD |
| XY_AR | 0.012930756 | 0.055530875 | 0.047659899 | 0.0203591239 | 0.11152909 | XR |

**Supplementary Table 4: The exact PG percentages within 100kpG and TOPMed show proximity to the gnomAD confidence intervals.**
